# Acellular Adipose Tissue promotes anti-fibrotic remodeling in Phase II Study

**DOI:** 10.64898/2026.02.04.26345214

**Authors:** Alexis N. Peña, Jordan A. Garcia, Amy E. Anderson, Joel C. Sunshine, Carisa M. Cooney, Pathik Aravind, Joseph Puthumana, Alexander T.F. Bell, Elana J. Fertig, Patrick Byrne, Damon S. Cooney, Jennifer H. Elisseeff

## Abstract

Acellular Adipose Tissue (AAT) is an off-the-shelf, cadaveric adipose-derived ECM-based biomaterial for soft tissue reconstruction. AAT has been validated preclinically to promote angiogenesis and adipogenesis and demonstrated safety, biocompatibility, and tolerability in a Phase I study. In this study we report the findings for the first ten patients in the Phase II study for permanent reconstruction of modest soft tissue defects. AAT promoted macrophages, CD3^+^ T cells, and CD34^+^ progenitor activity. Multiplex immunofluorescence staining using the PhenoCycler (formerly CODEX) imaging platform found that AAT can induce tertiary lymphoid structures (TLS). Nanostring GEOMx spatial transcriptional data analysis found significant differential gene expression between neighboring tissues with *EGR1*, *MCL1*, and *NR4A1* upregulated in AAT. These genes have roles in angiogenesis, anti-apoptotic processes, and promotion of anti-inflammatory genes, respectively. AAT promoted anti-fibrotic CD74^+^ adipose-derived stromal cells, confirmed by immunofluorescence staining. Our findings demonstrate that AAT promotes angiogenesis, adipogenesis, and anti-fibrotic remodeling.

## Background

The need for soft tissue reconstruction can arise from multiple causes such as congenital abnormalities, trauma, cancer resection, or aesthetic indications. Surgical options for soft tissue volume loss include local tissue transfer that often use adipose tissue. Regenerative medicine strategies aim to provide biological tissue replacements that mimic the native tissue. A number of these strategies are designed to functionally restore tissues by recruiting endogenous progenitor cells and immune cells to induce reparative processes. Just as important as promoting new tissue development, however, is inhibiting adverse outcomes like fibrosis. When treating soft tissue defects by filling with an injectable or implant, the host response can either support material integration and long-term function or mediate rejection, loss of function, or fibrosis. Induction of a foreign body response (FBR) characterized by fibrotic encapsulation of the implanted material with dense linearly aligned collagen deposition, chronic inflammation, and vascularization can be associated with poor aesthetic and functional outcomes for patients.

Currently there are no established regenerative medicine approaches for large soft tissue defects. Treatment options for large soft tissue defects include free, non-vascularized tissue transfer like autologous fat grafting, pedicled locoregional tissue transfer, and vascularized free flap procedures. Autologous fat grafting is an established clinical mainstay for aesthetic and reconstructive procedures where fat from a patient’s donor site is acquired generally by lipo-aspiration, washed, and then transferred to the defect site. Challenges associated with the technique include the need for multiple fat grafting sessions, potential donor site morbidity, unpredictable fat resorption (as much as 50%), necrosis, and potential persistence of calcifications and oil cysts [1, 2]. For larger soft tissue defects, non-vascularized fat grafts become limited by inadequate vascular supply. In such settings, vascularized adipofascial grafts have been employed with success but require microsurgical expertise and increased operative times [3, 4]. Additionally, not all patients are suitable candidates for fat grafting because they lack adequate donor fat tissue or had previous fat grafting attempts resulting in high resorption. Alternatives to fat grafting include synthetic implants as well as injectables comprised of synthetic, dermal-, collagen-, or hybrid-based materials, although these options are not suitable for larger defects. While implants are permanent, they are limited by their inability to be easily personalized for individual patients [5, 6]. Injectables have the flexibility to address unique patient contouring needs but have limited durability. Widely accessible volume restoration of moderate-to-large soft tissue defects remains a significant clinical unmet need.

Acellular Adipose Tissue (AAT) is an off-the-shelf therapy for soft tissue reconstruction designed to mimic native fat grafting. Off-the-shelf tissue products provide the advantage of being readily available to reconstruct defects of various sizes typically with reduced operating times and in outpatient settings. AAT is made from allograft adipose tissue that has been processed mechanically and chemically to remove lipids and cell debris. The final product is a colloidal injectable product that is terminally sterilized prior to clinical use. The processing technique preserves the composition and rheological properties of adipose tissue [7, 8]. Adipose extracellular matrix is the sole component of AAT. AAT retains the mechanical and biological properties of adipose tissue. Proteomic analysis of AAT revealed that collagen was the primary component and identified unique matrisome or matrisome-associated proteins distinct to adipose tissue [8]. Matrisome components support cell growth, differentiation, and tissue architecture [9]. The material serves as a framework for new, permanent tissue development. Preclinical studies with AAT in small and large animal models have demonstrated its safety and inductive role in adipogenesis and vascularization. Clinical evaluation of AAT for soft tissue reconstruction is ongoing. A Phase I clinical trial (NCT02817984) investigated a single discrete bolus of AAT in healthy volunteers undergoing an elective tissue-removal procedure. The study demonstrated that AAT is safe, biocompatible, and tolerable over the 18-week evaluation period. There were no serious adverse events associated with the AAT and there was increased cell migration. The Phase I clinical study validated the tissue development potential based on histopathological assessment of AAT explants. The Phase I study highlighted the scaffolds’ role in adipogenesis, supporting cellular activity, and had an instructive role for the immune system demonstrated by the enrichment of anti-inflammatory CD163^+^ macrophages and CD4^+^ T cells in AAT when compared to control fat in flow cytometric studies.

Preclinical studies and the Phase I clinical trial demonstrated AAT’s safety profile, biocompatibility, and ability to promote a pro-regenerative response for tissue repair. In the present study we report the initial results of the Phase II clinical trial (NCT03544632), a dose-escalation study conducted at Johns Hopkins University School of Medicine for the first ten patients, where AAT (Aegeria Soft Tissue) was permanently placed in modest (approx. 5-30cc) soft tissue defects of the chest or trunk. Primary objectives of the Phase II clinical trial include safety and efficacy assessments of AAT. Safety was determined by incidence and rate of adverse/unanticipated events. Efficacy will be evaluated at the completion of the study with additional patients. Secondary objectives include histopathology assessments of longitudinal core-needle biopsies and tolerability of AAT injections based on patient-reported satisfaction and physician-reported ease-of-use with the intervention surveys.

We performed an in-depth characterization of the host response to permanently placed AAT with core-needle biopsies in patients with soft tissue defects of the trunk. We used multiple imaging modalities to evaluate AAT. Histopathological analysis and immunofluorescence staining revealed increased cell migration and remodeling over time. Hematoxylin and eosin (H&E) staining demonstrated increased adipose tissue interspersed through the matrix thereby supporting adipogenesis and new tissue development. Progenitor activity was confirmed by CD34 immunofluorescence staining. Immunofluorescence– and H&E-guided Nanostring GEOMx spatial transcriptional data from AAT biopsies revealed unique cell composition and gene expression based on tissue type (AAT, proximal scar, and proximal fat), with AAT promoting genes related to angiogenesis, phagocytic activity, and an adaptive immune response. An anti-fibrotic CD74^+^ stromal cell subpopulation was abundant in the injected AAT material. Patient and physician surveys were taken at multiple time points over the one year follow up and both scored AAT injections highly for “naturalness”, supporting the tissue-like properties of AAT and tissue development. The participant surveys and pathology report also validated tolerability of the material in concordance with Phase I clinical findings.

### Results

#### Trial design

Ten patients (7 females, 3 males; aged 20-65; average age 49; Supplemental Table 1 and. 2) were enrolled in an IRB– and OHRO– (formerly known as the HRPO)-approved Phase II clinical trial (NCT03544632) (Supplemental Figure 1). We report on the data from the first 10 patients who completed the study, including the 1-year follow-up after initial injection. Patient inclusion criteria included 18-65 years age range with no history of diabetes or cancer-related care in the two years prior to enrollment. Patients were limited to those with modest soft tissue defects of the chest or torso. Over 2,400 patients were screened for enrollment by medical record review, telephone, and/or physical examinations to assess appropriateness of the defects. For enrolled patients, the trial’s dose-escalation design allowed for 4-40 cc of material to be injected, starting with either 4 or 10 cc or 20 cc. After the Month 3 follow-up visit, participants had the option to undergo reinjection (not to exceed a total injection amount of 40 cc) if the defect volume was not adequately restored and the participant did not experience any adverse events that would contraindicate an additional AAT injection. Three patients elected to receive a second injection ranging from 8-20 cc of additional material. Six defects were of the chest wall, while four were located on the abdominal wall (Supplemental Table 3). Three patients had a history of radiation.

#### Permanently placed AAT is safe and tolerable in patients with modest soft tissue defects of the trunk

The overall goal of the clinical testing program is to evaluate modest volume (approx. 5-30cc) reconstruction of the trunk using AAT. The first ten patients were enrolled and treated with either 4 cc or 10 cc of AAT material, depending on the participant’s assigned treatment group, via sterile subcutaneous injection into the target defect. Safety was assessed by serial clinical examinations and laboratory studies for one year after the initial injection as outlined in the study protocol. An overview of physical exam findings can be found in Supplemental Table 4. For the first ten patients in the study, no patients experienced serious adverse events in concordance with the Phase I trial [8]. Three patients experienced self-resolving hyperpigmentation or ecchymosis at the injection site. No patients demonstrated significant inflammation, redness, swelling, or pain at the injection sites. Summaries of adverse events are shown in Supplemental Table 5-6.

Patient-reported satisfaction surveys demonstrated that, overall, AAT was well tolerated. During follow up visits, participant and physician surveys were provided to evaluate fullness at injection site, softness, smoothness, and naturalness of implants. Participants rated comfort during and 30 minutes after the injection and the physician reported ease of use and overall appearance. Subject– and investigator-reported appearance scores were based on softness, smoothness, and naturalness, using a 5-category scale 1 being ‘very hard’, ‘very lumpy’, or ‘very unnatural’ to 5 being ‘very soft’, ‘very smooth’, or ‘very natural’, respectively. The satisfaction score was based on three scores: comfort during injection, comfort 30 minutes after injection, and fullness of injected site. All used a 5-category scale with 1 being ‘very dissatisfied’ to 5 being ‘very satisfied’. The summary of the results is in Supplemental Tables 7-9. The satisfaction rate among participant and physician surveys was determined by the percentage of scores that were rated 4 or 5. Based on participant surveys, the abdominal cohort more often reported satisfactory comfort of injection both during and 30 minutes after injection compared to the chest cohort (Supplemental Figure 2 and 3). The chest cohort less often reported comfort during injection (17%) and satisfaction was restored 30 minutes after injection (100%). Participants from both cohorts reported satisfaction with the naturalness of the injection. The chest cohort more often reported satisfactory fullness of the injection site than the abdominal cohort. Overall, both cohorts reported satisfaction with the smoothness of the injection. The abdominal cohort reported some dissatisfaction in smoothness during the first month post-injection which improved thereafter. For both cohorts, there was improved satisfaction with softness of injection over the course of follow-up visits. The physician reported overall satisfaction with ease of use of the product, reporting superior satisfaction when used in the abdominal cohort (100%) compared to the chest cohort (67%) (Supplemental Figure 4-5). While the physician reported some dissatisfaction with the fullness of the injection site, particularly in the abdominal cohort, overall physician-reported satisfaction with AAT naturalness and softness was positive. Finally, while the physician reported lower satisfaction with fullness in abdominal defects (25%), 80% of all the participants were satisfied with the fullness of their injection sites one year post-injection.

Given AAT’s allogeneic origin, panel reactive antibody (PRA) testing was performed at baseline prior to AAT injection, and at 4 weeks and 12 months post-injection. PRA testing is standard for detection of anti-human leukocyte antigen (anti-HLA) antibodies. From the Phase I data, we have previously reported that patients did not develop new anti-HLA antibodies up to 12 weeks post-injection, supporting the finding that AAT is biocompatible [8]. For the first ten patients in the Phase II study, none of the patients enrolled had increased HLA antibodies at 4 weeks and 12-months post-injection. There were no patients that developed new anti-HLA antibodies post-AAT injection. These additional data at longer timepoints support the claim that AAT does not cause a systemic immunologic response. PRA testing results are summarized in Supplemental Table 10.

#### AAT *in situ* can be visualized with various imaging modalities and ultrasound can guide optimal biopsy acquisition

Multiple imaging modalities were investigated to image the soft tissue defects and to detect the presence of AAT material over time: 2D photography, 3D photography, computed tomography (CT), and ultrasound (US). Standardized 2D and 3D photography were performed to document soft tissue defect location and volume maintenance over time (Fig. 1a).

**Figure 1.**
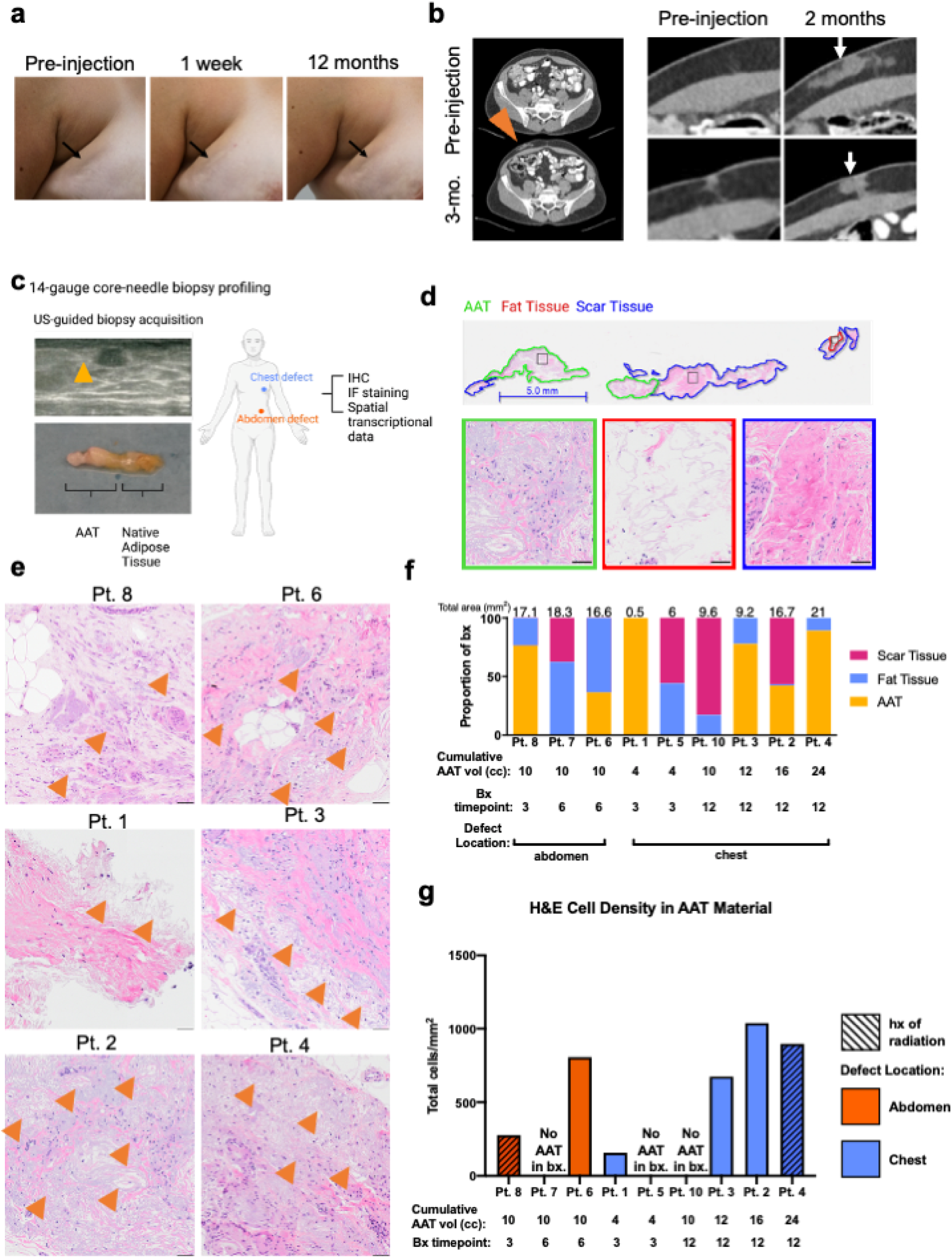
AAT *in situ* can be visualized with various imaging modalities and ultrasound can guide optimal core-needle biopsy acquisition. **a.** 2D photography of soft tissue defect at pre-injection and 1 week and 12 months post-initial injection **b.** Axial CT scans pre-injection and 8 weeks post-injection with arrows pointing to AAT material **c.** Ultrasound guided 14-gauge core-needle biopsy acquisition with ultrasound images. Arrow pointing to AAT material. Gross image of core needle biopsy containing AAT and Native Adipose Tissue. Core-needle biopsy characterization strategy using IHC, IF, and spatial bulk genomic data **d.** Representative whole slide H&E of core-needle biopsy section annotated by tissue type. H&E scale bar=5 mm, zoom-in scale bar=50 µm **e.** H&E images of AAT material in core-needle biopsy by patient. Scale bar=100 µm **f.** Bar plot of proportion of core-needle biopsy of H&E data by tissue type with total area analyzed highlighted **g.** H&E Cell density quantification in AAT Material quantified using HALO

In a few patients, CT scans performed as a part of patients’ standard care (e.g. not for study purposes) included the AAT material. When available, standard care CT scans were used to assess the shape and structure of the injected material, which appeared hyperintense relative to surrounding adipose tissue (Fig. 1b). The CT scans could also accurately estimate AAT volumes, although it could not differentiate newly formed tissue from the native surrounding tissue (Supplemental Table 11). It is important to note the appearance of the AAT in CT, MRI, and other modalities so that radiologists can differentiate from abnormal tissue lesions.

AAT material can also be identified with ultrasound (US). Injected material appears hypoechoic on ultrasound relative to surrounding tissue (Fig. 1c). Ultrasonography was used to guide CORE needle biopsies to maximize the amount of AAT in each biopsy. An example of a gross image of core-needle biopsy with AAT material and native adipose tissue is shown in Fig. 1c. Core-needle biopsies were acquired for secondary endpoints related to histopathological assessments including immunohistochemistry (IHC), immunofluorescence staining, and Nanostring GEOMx spatial transcriptional data analysis. One core-needle biopsy per patient was performed at one of four study time points: 3-, 6-, or 12-mo. post-initial injection. Biopsies were acquired to assess presence of AAT and the cellular response to the material. Some biopsies did not survive the tissue processing steps of fixing and embedding in paraffin wax. In total, 6 specimens were suitable for further histological analysis (see Supplemental Figure 1 for clinical study design). The core-needle biopsy acquisition technique captured, to a varying amount, AAT and surrounding tissue which included proximal fat tissue, scar tissue, or both. To investigate the host response to AAT injections, biopsy regions were analyzed by tissue type and the total area of FFPE sections analyzed were reported (Fig. 1d-f).

#### AAT promotes cell infiltration and remodeling over time

To assess appearance of tolerability histologically and to evaluate cell migration and material evolution *in situ*, H&E staining was performed on core-needle biopsies. Histopathological assessment of core-needle biopsies by a trained pathologist found that there was cellular infiltration into the AAT material, remodeling, and that AAT appeared well tolerated. A semiquantitative histologic scoring system (Table 1) was used to evaluate cell type and presence. Scores for individual biopsy samples from the pathology report are included in Table 2. Histologic examination demonstrated that cellular and tissue response to the implant increased with time and was associated with degradation and remodeling of the acellular substrate when present. This remodeling was accompanied by chronic inflammation, with lymphocytes and giant cells present in most biopsies. Fat necrosis and acute inflammatory cells were minimal or absent. No polymorphonuclear cells were identified by H&E. While remodeling did occur, notably, AAT material was still present in the 12-month biopsies and appeared randomly organized. A fibrosis response was generally minimal or not detected in biopsies. Fibrosis, defined as a histopathologic descriptor of the thickening and scarring of connective tissue, which histologically is represented by zones of normal tissue replaced by bands of increased fibroblasts in a sclerotic stroma, were minimal in 5 biopsies and mild in 1 biopsy. AAT remodeling was accompanied by neovascularization as well as possible adipogenesis, as evidenced by adipocyte presence throughout the material (Fig. 1e). To quantify the cell density in the biopsies, 20x whole-slide images of the H&E biopsies were acquired, annotated by tissue type, and analyzed in HALO, the gold standard quantitative image analysis platform (RRID:SCR_018350). There were increased cell densities in the AAT regions generally by biopsy timepoint (Fig. 1g). Cell density in fat regions of the biopsies had more comparable cell densities between patient biopsies, apart from patient 3 and 2 which had higher cell densities (Supplemental Figure 9a). When scar was present in biopsies, the cell density was variable (Supplemental Figure 10a).

**Table 1.** Histological Examination Scoring System.

| Cell Type/Response | Score |  |  |  |  |
| --- | --- | --- | --- | --- | --- |
|  | 0 | 1 | 2 | 3 | 4 |
| Polymorphonuclear cells | 0 | Rare, <25/hpf | 25-50/hpf | 51-75/hpf | Packed |
| Lymphocytes/histiocytes | 0 | Rare, <25/hpf | 25-50/hpf | 51-75/hpf | Packed |
| Giant cells | 0 | Rare, <10/hpf | 11-20/hpf | 21-30/hpf | 31-40/hpf |
| Congestion | 0 | Minimal | Mild | Moderate | Severe |
| Mineralization | 0 | Minimal | Mild | Moderate | Severe |
| Fibrosis | 0 | Minimal | Mild | Moderate | Severe |
| Edema fluid accumulation | 0 | Minimal | Mild | Moderate | Severe |
| Anisotropic Material | 0 | Minimal | Mild | Moderate | Severe |

Note: hpf = high-powered (400X) field

**Table 2.** Histopathological Examination Scoring System—Results (adapted from official pathology report; ordered by defect location and cumulative AAT volume)

| Cell Type/Response | Case Number by defect location |  |  |  |  |  |  |  |  |
| --- | --- | --- | --- | --- | --- | --- | --- | --- | --- |
|  | Abdomen cohort |  |  | Chest cohort |  |  |  |  |  |
|  | 8 | 7 | 6 | 1 | 5 | 10 | 3 | 2 | 4 |
| Polymorphonuclear cells | 0 | 0 | 0 | 0 | 0 | 0 | 0 | 0 | 0 |
| Lymphocytes/histiocytes | 2 | 1 | 1 | 1 | 1 | 1 | 1 | 2 | 2 |
| Giant cells | 1 | 0 | 1 | 0 | 0 | 0 | 1 | 2 | 2 |
| Congestion | 0 | 0 | 0 | 0 | 0 | 0 | 0 | 0 | 0 |
| Mineralization | 0 | 0 | 0 | 0 | 0 | 0 | 0 | 0 | 0 |
| Fibrosis | 1 | 1 | 1 | 0 | 0 | 0 | 2 | 1 | 1 |
| Edema fluid accumulation | 0 | 0 | 0 | 0 | 0 | 0 | 0 | 0 | 0 |
| Anisotropic Material | 0 | 0 | 0 | 0 | 0 | 0 | 0 | 0 | 0 |

#### Tertiary lymphoid structures are present around the AAT material

The immune system has an essential role in tissue repair processes. Biological scaffold implantation is known to modulate the tissue microenvironment and promote a pro-regenerative immune cell phenotype [8]. The previous investigation of AAT over 18 weeks *in situ* in the Phase I clinical study found CD4^+^ and CD8^+^ T cells clustered with other non-T cells suggesting TLS formation [8]. Preclinical models using biological scaffold implantation found that the scaffolds promoted clusters of adaptive immune cells. A murine model studying the impact of ECM scaffold treatment and tumor formation found that the ECM microenvironment altered cell recruitment and promoted localization of CD3^+^ T cells and B220^+^ B cells [10]. In a preclinical study investigating B cell changes after ECM treatment in a murine volumetric muscle loss model, ECM treatment induced germinal center formation in the draining lymph nodes. In that study, AAT core-needle biopsies were used to investigate clinical relevance to preclinical findings. Clusters of B cells were found in Phase II AAT core-needle biopsies [11]. To profile the adaptive immune response and antigen presentation in AAT core-needle biopsies, highly specific high-plex immunofluorescence phenotyping was performed with the PhenoCycler (formerly CODEX) imaging platform. There were organized clusters of CD3^+^CD4^+^ and CD3^+^CD8^+^ T cells and CD20^+^ B cells present in AAT core-needle biopsies. HLA-DR^+^ antigen presenting cells and CD31^+^ endothelial cells were also present in the cluster. These clusters suggest that AAT promotes tertiary lymphoid structures (TLS) (Fig. 2a-c). In the present study, CD20^+^Ki-67^+^ cells revealed actively proliferating B cells in the B cell clusters within the TLS. Proliferating B cells in TLS suggest the existence of germinal center-like B cells [12]. Tertiary lymphoid structures are organized lymphoid aggregates that can range from small discrete clusters of lymphocytes to larger structures resembling the organization of lymph nodes (Supplemental Figure 11a). TLS provide local immune cell niches in the microenvironment to promote adaptive immunity. In addition to proliferative B cells in AAT material, there were other Ki-67^+^ cells present demonstrating active cell proliferation (Supplemental Figure 11b).

**Figure 2.**
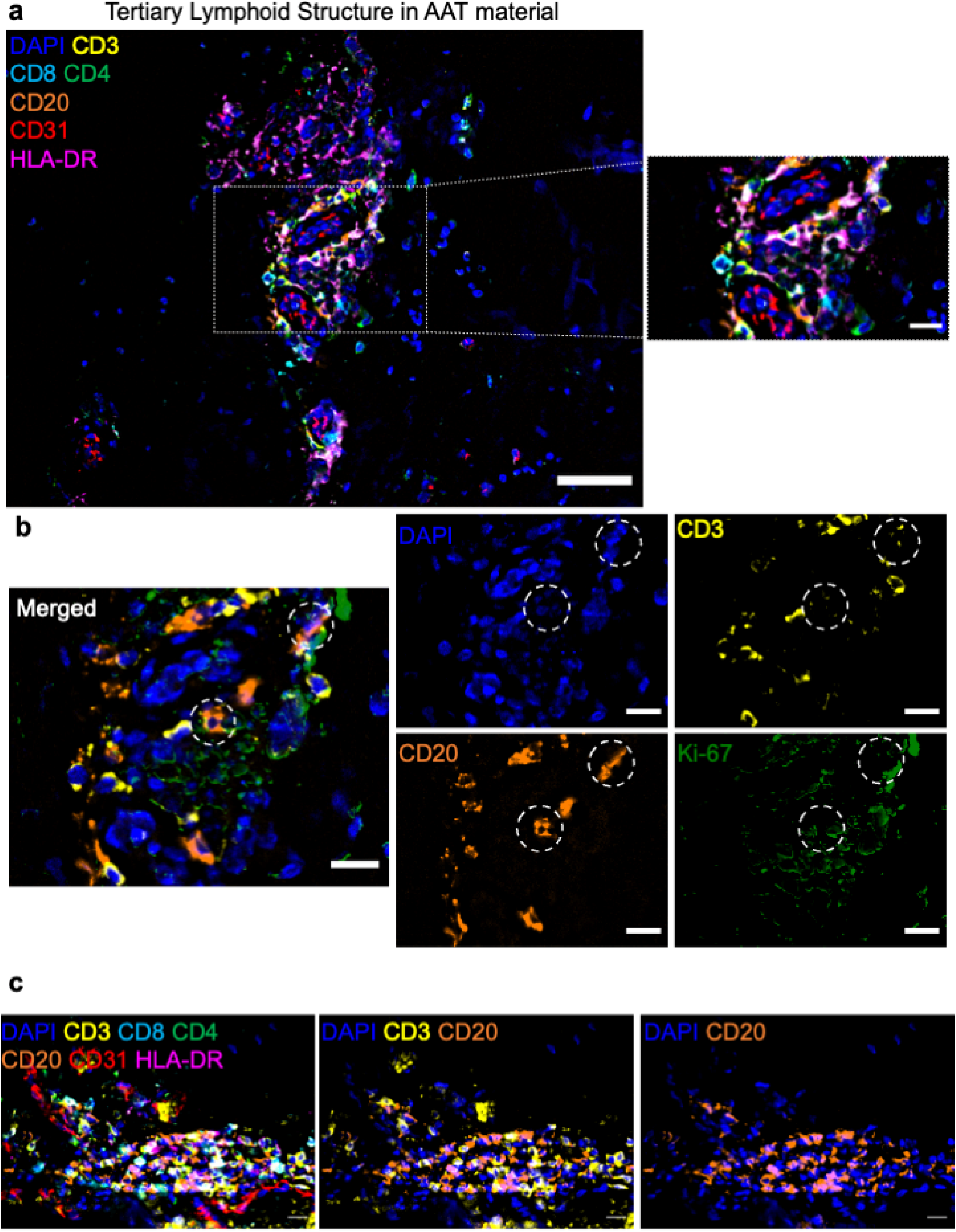
Tertiary lymphoid structure (TLS) in AAT core-needle biopsies. **a.** Multiplex immunofluorescence images using the PhenoCycler of AAT Material profiling blood vessels (CD31^+^), adaptive immune cells: B cells (CD20^+^), CD3^+^ T Cells (CD4^+^ and CD8^+^), and antigen presenting cells (APCs) (HLA-DR^+^) revealed tertiary lymphoid structures in AAT. Zoomed out image; scale bar=50 µm. Zoomed in image; scale bar=15 µm **b.** Multiplex immunofluorescence images reveal additional lymphoid aggregates in AAT material. Scale bar=15 µm **c.** Multiplex immunofluorescence images revealed Ki-67^+^CD20^+^ proliferative B cells in aggregate with CD3^+^ cells. Scale bar=15 µm

**Figure 3.**
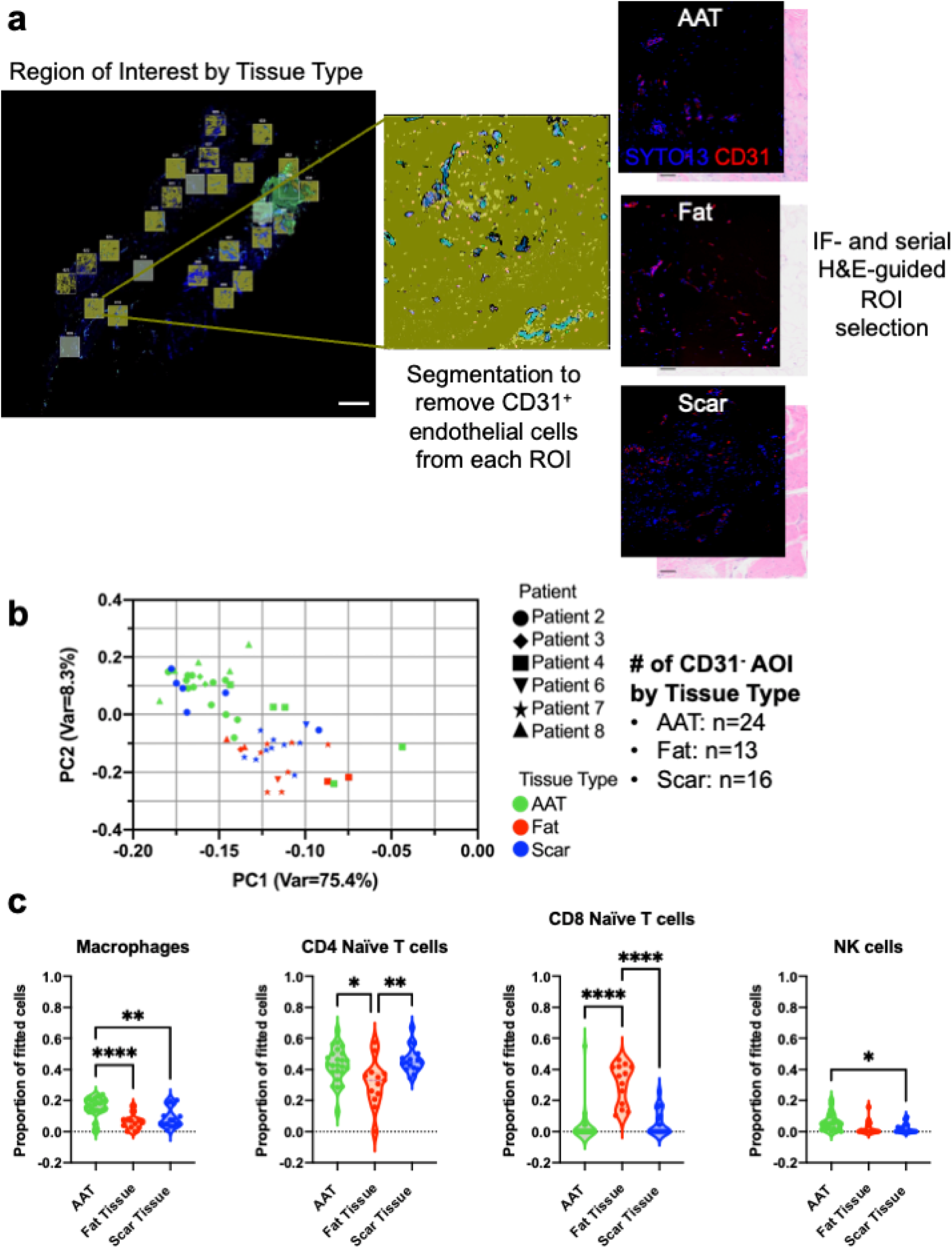
Spatial high-plex gene expression analysis of AAT core-needle biopsies. **a.** ROI selection strategy for Nanostring DSP gene expression analysis by tissue type using serial H&E and IF staining; SYTO13 (nuclear stain) and CD31 (endothelial cell marker). Scale bar=1 mm; Each ROI is approximately 647 x 718 µm. Each ROI was segments to remove CD31^+^ endothelial cells **b.** PCA of CD31^-^ AOI. Nanostring DSP gene expression data colored by tissue type **c.** Spatial cell deconvolution results by tissue type using SpatialDecon algorithm revealed distinct cell type composition by tissue type. Ordinary one-way ANOVA with multiple comparisons p <0.05

#### TLS-associated immune and stromal cell composition in AAT injections

To study the cell composition and gene expression in different tissue types within the biopsies, Nanostring GEOMx Digital Spatial Profiler (DSP) transcriptional data analysis was employed. The classic marker for endothelial cells and their progenitors (CD31) and nuclear stain (SYTO13) and serial H&E were used to guide precise region of interest (ROI) selection. ROI were selected to ensure one tissue type (AAT, fat, scar) per ROI (Fig. 3a). Spatially resolved gene expression data from 6 patients focused on the CD31^-^ segmented AAT ROI (n=24), fat ROI (n=13) and scar ROI (n=16). The hierarchical clustering of the CD31^-^ Nanostring DSP data do not cluster completely by tissue type, biopsy timepoint or defect location (Supplemental Figure 12a). The principal component analysis (PCA) showed that PC1 stratifies AOIs (segmented ROI) by tissue type (Fig 3b). There was overlap between fat and scar AOIs and as well as AAT and scar AOIs, although fat and scar AOIs had more overlap in the PCA space.

Cell composition estimates were quantified from the Nanostring GEOMx spatial transcriptional data using the SpatialDecon algorithm. The SpatialDecon algorithm is a cell deconvolution technique that uses reference single cell RNA sequencing (scRNA-seq) cell profiles to map cell type estimates in spatial gene expression studies [13]. In this case, the reference scRNA-seq data was the ImmuneTumor_safeTME normalized cell matrix used as the default in Nanostring’s analysis pipeline. Based on the SpatialDecon results for the CD31^-^ AOI, there were unique cell type composition dependent on tissue type. Macrophages and naïve CD4^+^ T cells were the most abundant cell types in AAT (Supplemental Figure 12b). These findings are in concordance with flow cytometry results from the Phase I explants [8]. There were significantly more macrophages in AAT compared to proximal scar and fat (Fig. 3c). NK cells were significantly more abundant in AAT compared to proximal scar. Naïve CD8^+^ T cells and naïve CD4^+^ T cells were the most abundant cell types in proximal fat, with there being significantly more naïve CD8^+^ T cells in fat compared to AAT and scar. Naïve CD4^+^ T cells and plasma cells were most abundant in proximal scar (Supplemental Figure 12b). These findings demonstrate that AAT recruits unique cell types regardless of surrounding tissue.

#### Distinct differential gene expression in AAT compared to neighboring tissues

We next examined gene expression differences within the core-needle biopsies. Gene expression analysis of Nanostring GEOMx DSP CD31^-^ AOI comparisons by tissue type revealed differentially regulated genes (p_adj_ < 0.05, 0.58 ≤ LFC ≤ –0.58). Comparison of AAT material and proximal fat found 49 significantly differentially upregulated genes in AAT and 56 genes upregulated in fat (Fig. 4a). There were various macrophage-related genes upregulated in AAT (*CHIT1, HLA-DRA, HLA-DRB4, HLA-DRB3, HLA-DQA1, CD68, CD74*). The macrophage migration inhibitory factor (MIF)/CD74 signaling pathway is strongly implicated in inflammation, angiogenesis, and protective roles during injury [14, 15]. *SPP1* is part of the integrin alphav-beta3 complex. The adhesion receptor integrin αvβ3 is a marker of angiogenic vascular tissue [16]. Integrin alphax-beta2 complex was represented by *ITGB2* and *ITGAX.* The complex is important for leukocyte trafficking and immunological processes. Various differentially upregulated genes in fat were related to adipocyte proliferation, differentiation, energy balance, and lipid transport (*CEBPA, PRKAR2B, LEP, CD36*). We used STRING to construct a protein-protein interaction (PPI) network of the differentially regulated genes in AAT (Supplemental Figure 12d) [17]. To detect functional enrichment, overrepresentation tests were performed using STRING. Using the Gene ontology annotations for biological process, molecular function, and cellular component as functional pathway classification frameworks, there was functional enrichment of *Positive regulation of monocyte differentiation* (GO:0045657) (*CD74, CD4, JUN*); FDR=0.00043, *MHC class II receptor activity* (GO:0032395) (*HLA-DPA1, HLA-DQB1, HLA-DQA1l, HLA-DRA*); FDR=1.84e-5, and *Transcription factor AP-1 complex* (GO:0035976) (*JUN, JUNB, FOS*); FDR=1.71e-5. Enrichment detection using REACTOME Pathways found adaptive immune related pathways enriched such as *Phosphorylation of CD3 and TCR zeta chains* (HSA-202427) (*PTPRC, CD4, HLA-DPA1, HLA-DQB1, HLA-DRB5, HLA-DRA*); FDR=9.18e-9 and *Translocation of ZAP-70 to immunological synapse* (HSA-202430) (*CD4, HLA-DPA1, HLA-DQB1, HLA-DRB5, HLA-DRA*); FDR=3.6e-7. Automated pathway enrichment analysis of the PPI network constructed from DEG in fat using the Biological process category of Gene Ontology found enrichment of *vascular wound healing* (GO:0061042) (*GATA2, VEGFA, CD34, MCAM);* FDR=6E-6 (Supplemental Figure 12e). Using REACTOME Pathways there was functional enrichment of *VEGF binds to VEGFR leading to receptor dimerization* (HSA-195399) (*VEGFA, VEGFB*); FDR=0.0270. Using subcellular localization (COMPARTMENTS) there was functional enrichment of *VEGF-A complex* (*VEGFA, VEGFB*); FDR=0.0270 and *Plasma lipoprotein particle* (*LEP, SAA1, CLU*); FDR=0.0317.*VEGF* can stimulate angiogenesis and is an anti-apoptotic factor [18].

**Figure 4.**
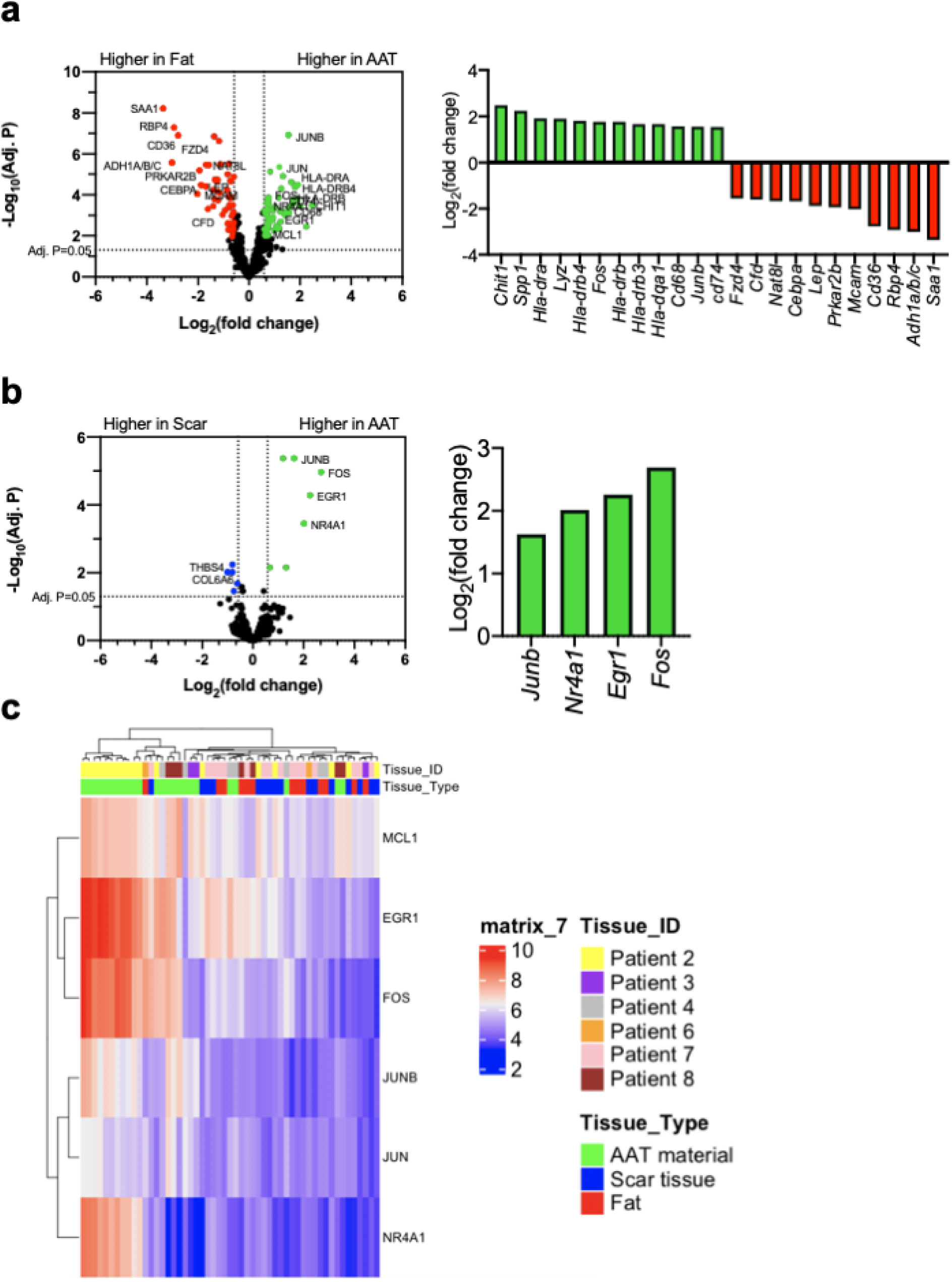
Spatial high-plex gene expression analysis of AAT core-needle biopsies. **a.** Volcano plot of differentially expressed genes comparing AAT and fat in core-needle biopsies revealed macrophage-related genes upregulated in AAT (*Chit1, Hla-dra, Hla-drb4, Hla-drb3, Hla-dqa1, Cd68, Cd74*) and genes associated with the activation of AP-1 transcription factor (*Fos, Jun, Junb)*. Various differentially upregulated genes in fat were associated with adipocyte proliferation, differentiation, energy balance, and lipid transport (*Cebpa, Prkar2b, Lep, Cd36*). Linear mixed model (LMM) test using Limma, design accounts for individual patients, p_adj_ < 0.05, 0.58 ≤ LFC ≤ –0.58 **b.** Volcano plot of differentially expressed genes comparing AAT and scar in core-needle biopsies. Genes upregulated in AAT were related to AP-1 transcription factor. Linear mixed model (LMM) test using Limma, design accounts for individual patients, p_adj_ < 0.05, 0.58 ≤ LFC ≤ –0.58 **c.** Heatmap highlighting genes differentially upregulated in AAT compared to both fat and scar, that *Egr1, Fos, Jun, Junb, Mcl1,* and *Nr4a1.* Row values are vst normalized gene counts.

Comparison of AAT material and proximal scar found 7 significantly differentially upregulated genes in AAT and 5 genes upregulated in scar (Fig. 4b). Enrichment analysis of the PPI-network constructed from differentially expressed genes in AAT found significant enrichment of terms and pathways related to transcription factor AP-1 (Supplemental Figure 12f). Using Gene Ontology Category Cellular Component there was function enrichment of *Transcription factor AP-1 complex* (GO:0035976) (*JUN, JUNB, FOS*); FDR=1.66e-6. Using REACTOME Pathways there was functional enrichment of *Activation of AP-1 family of transcription factors* (HSA-450341) (*FOS, JUN*); FDR=0.0032. Comparisons between fat and scar were also analyzed; with 24 genes being differentially upregulated in fat and 8 in scar (Supplemental Figure 12c). PPI constructed from DEG in fat had functional enrichment of pathways and terms related to adipose biology (Supplemental Figure 12g). Biological process category from Gene ontology found function enrichment of *Lipid export from cell* (GO:0140353) (*LEP*); FDR=0.0155. Using REACTOME Pathways there was functional enrichment of *Transcriptional regulation of white adipocyte differentiation* (HSA-381340) (*SREBF1, LEP, CEBPA*); FDR=0.00012. Using WikiPathways there was functional enrichment of *Transcriptional cascade regulating adipogenesis* (WP4211) (*SREBF1, CEBPA*); FDR=0.0310.

PPI constructed of DEG in scar had functional enrichment of terms related to ECM and cell adhesion: REACTOME Pathways found enrichment of *Extracellular matrix organization* (HSA-1474244) (*FN1, LTBP1*); FDR=0.0084 and annotated Keywords (UniProt) had functional enrichment of *Cell adhesion* (KW-0130) (*CD9, FN1, FAP*); FDR=0.00031 (Supplemental Figure 12h). There were genes that were significantly differentially upregulated in AAT compared to both fat and scar regions that include *EGR1, FOS, JUN, JUNB, MCL1,* and *NR4A1* (Fig. 4c). *EGR1* is important in angiogenesis, hemopoiesis, and regulating genes related to ECM. *MCL1* is an anti-apoptotic protein for cell survival. Orphan nuclear hormone receptor *NR4A1* regulates pro-inflammatory phenotype and is essential to Ly-6C^low^ monocyte production (reparative monocytes). *NR4A1* can directly induce expression of anti-inflammatory genes [19]. AP-1 transcription factor are regulators of cell proliferation, differentiation, and transformation. AP-1 transcription factor complex can influence adipocyte commitment [20].

ECM scaffold treatments are known to modulate the tissue microenvironment to recruit pro-regenerative innate and adaptive immune cells [10, 11, 21, 22]. Altogether, the gene expression analysis demonstrates that AAT influences distinct cell activity compared to proximal fat and scar. There were genes upregulated in AAT related to phagocytic activity that is important in tissue remodeling. The PPI functional enrichment analysis identified pathways and terms related to angiogenesis, phagocytosis, cell adhesion and trafficking, and adaptive immune response in AAT. The transcription factor AP-1 complex was also significantly differentially upregulated in AAT compared to both fat and scar suggesting promotion of pre-adipocyte differentiation [23].

#### Macrophage polarization in AAT biopsies

Macrophages are critical for wound healing and exist on a spectrum of diverse macrophage phenotypes ranging from pro-inflammatory classically denoted as M1 macrophages to anti-inflammatory generally regarded as alternatively activated M2 macrophages. Biological scaffolds promote a pro-regenerative M2 macrophage phenotype. Previous flow cytometric investigation of cell infiltration and phenotype within *in situ* AAT (excision points at 1 to 6 weeks) found an enhanced hybrid M1/M2 macrophage response with most macrophages being alternatively activated M2 macrophages and a large proportion of double positive macrophages (CD163^+^CD80^+^). In the Phase I study, M1 macrophages were the least abundant macrophage population in AAT explants [21]. To better understand the macrophage phenotype in core-needle biopsies, immunofluorescence triple staining of pro-inflammatory CD80 and mannose receptor C-type 1 MRC1/CD206 with pan-macrophage CD68 marker were performed (Fig. 5a, Supplemental Figure 13a). The proportion of biopsy by tissue type and total area analyzed were shown in Supplemental Figure 13b. Cell densities from whole slide images for CD68^+^CD80^-^CD206^-^, CD68^+^CD80^+^CD206^-^, CD68^+^CD206^+^CD80^-^, and CD68^+^CD80^+^CD206^+^ were calculated using HALO. CD68^+^ macrophages were found in all biopsies with varying densities (Fig. 5b). Most CD68^+^ macrophages were unpolarized (double-negative), 3 of the 4 biopsies stained (Pt. 8, Pt. 3, Pt. 2) had more CD68^+^CD206^+^CD80^-^ cells than CD68^+^CD80^+^CD206^-^ cells (Fig. 5c). Biopsies that had more CD68^+^CD80^+^CD206^-^ cells tended to have more double-positive cells (Pt. 2 and Pt. 4). Proximal fat tissue had fewer CD68^+^ cells than in AAT (Supplemental Figure 9b) and generally fewer double-negative macrophages except for Pt. 5. In general, the proportion of CD68^+^CD206^+^CD80^-^ cells were more abundant in fat tissue. The double positive population made up 25% of the macrophage population in 2 of the 5 biopsies that had fat tissue (Pt. 3 and Pt. 4). CD68^+^ cells in scar tissue were primarily double-negative and most were CD68^+^CD206^+^CD80^-^ cells; there were few CD68^+^CD80^+^CD206^-^ cells and double-positive cells (Supplemental Figure 10b-c).

**Figure 5.**
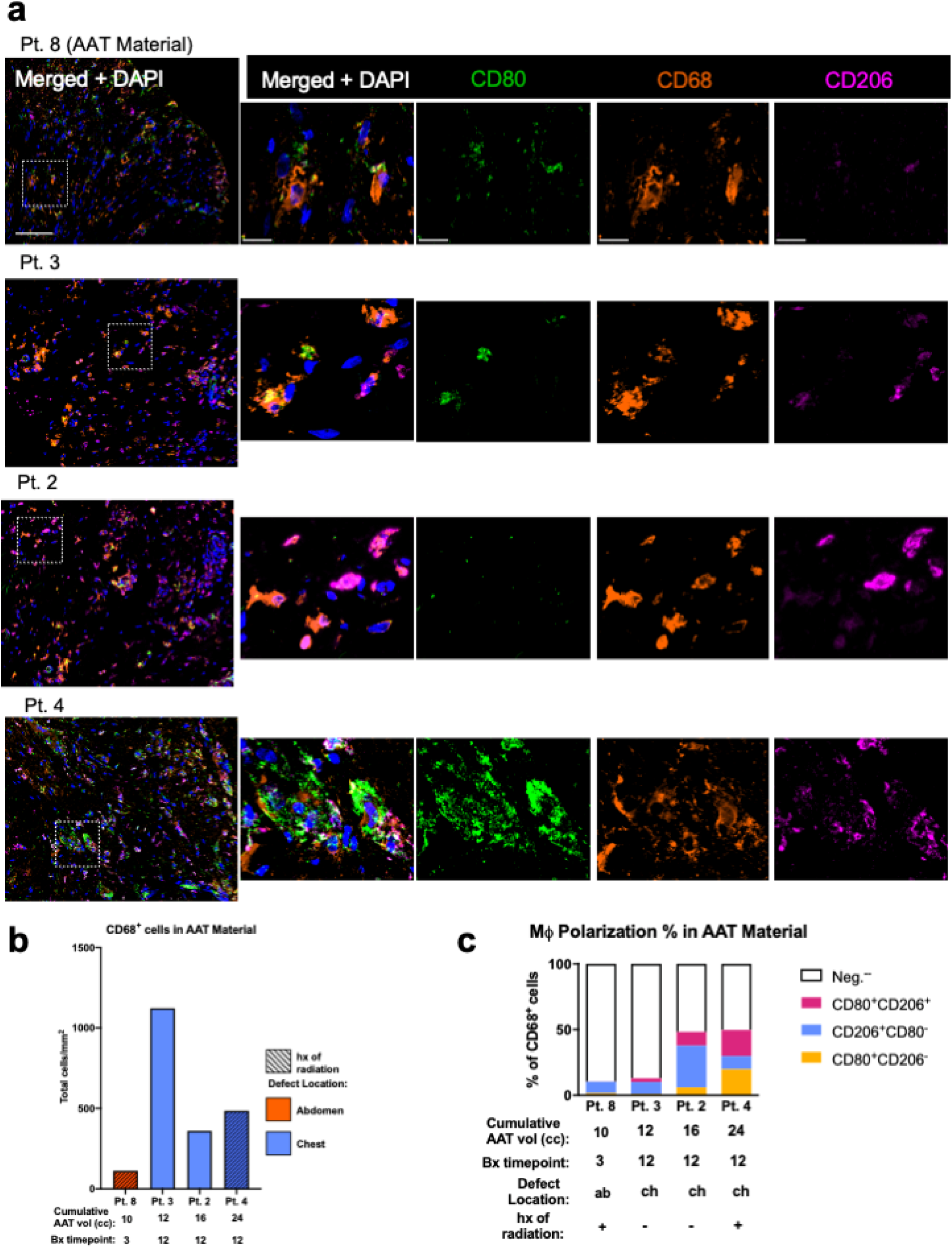
Macrophage polarization of AAT core-needle biopsies. **a.** Multiplex immunofluorescence images of AAT to investigate macrophage (CD68^+^) phenotype using classically-activated pro-inflammatory “M1” CD80^+^ and alternatively activated anti-inflammatory “M2” CD206^+^ markers. Zoomed out image; scale bar=100 µm. Zoomed in image; scale bar=20 µm **b.** CD68^+^ cell density in AAT material quantified by IF staining using HALO **c.** Macrophage polarization in AAT material quantified by IF staining using HALO. Populations of nonpolarized (CD68^+^CD80^-^CD206^-^ cells), classically-activated (CD68^+^CD80^+^CD206^-^ cells), alternatively activated (CD68^+^CD206^+^CD80^-^ cells), and double-polarized (CD68^+^CD80^+^CD206^+^ cells) represented as a percentage of CD68^+^ cells

Cellular senescence is a response characterized by stable cell cycle arrest that can be induced by a variety of stimuli not limited to DNA damage, oxidative stress, and oncogene activation. Senescent cells contribute to the tissue microenvironment through the production of senescence-associated secretory phenotype (SASP) factors. Although the prolonged presence of senescent cells can cause deleterious outcomes, there is an essential role for transient senescent cells in optimal wound healing. There are some beneficial roles for SASP in tissue repair particularly in older individuals whereby certain SASP factors can reduce fibrosis [24–26]. SASP factors can also recruit macrophages and influence macrophage polarization. Vascularization of tissue engineering scaffolds is a coordinated effort between multiple macrophage phenotypes [27]. To investigate the senescent status and the spatial relationship between macrophages and blood vessels present in AAT, immunofluorescence staining of senescent marker tumor suppressor p16, pan-macrophage marker CD68 and endothelial cell marker CD31 was performed (Supplemental Figure 14a, Supplemental Figure 15a). The proportion of biopsy by tissue type and total area analyzed were shown in Supplemental Figure 14b. There was an abundance of macrophages and giant cells near blood vessels in AAT regions. p16^+^ cells were present in AAT at small densities ranging from 3-24 cells/mm^2^ (Supplemental Figure 14b). Even fewer p16^+^ cells in fat tissue (0-0.4 cells/mm^2^) (Supplemental Figure 16a). p16^+^ cells were more abundant in scar tissue when present at 198 cells/mm^2^ (Supplemental Figure 16b). There were p16^+^ macrophages and endothelial cells in AAT and scar but not in fat tissue, although the majority of p16^+^ cells were CD68^-^ and CD31^-^ (Supplemental Figure 16a-b).

#### AAT promotes antifibrotic effects and tissue remodeling

To examine AAT collagen network remodeling by infiltrating cells, we performed Picrosirius red (PSR) staining. Picrosirius red is a standard dye for analyzing collagen organization that enhances collagen’s natural birefringence. When imaged under polarized light, the color and intensity of the birefringence from collagen fiber changes dependent on amount of fibril packing or collagen thickness [28, 29]. Thicker, closely packed collagen fibrils show up as strong red or yellow birefringence, while thinner, loosely packed fibers show up as weak green birefringence. Scar tissue is generally characterized by thick, dense linearly organized collagens. Picrosirius red staining showed thicker collagen matrix in AAT regions for Pt. 8 (abdomen cohort, 3-mo. biopsy). At interfaces with native tissue and cell infiltrate into AAT there were distinct remodeled AAT regions that have thinner more green collagen fibers and areas with less cell infiltrate that were redder with thicker collagen (Supplemental Figure 17a). AAT regions in the other biopsies (Pt. 3, Pt. 2, Pt. 4, chest cohort, 12-mo. after initial injection biopsy) had less organized, less abundant, and thinner green collagen fibers (Fig. 6a). Overall, the collagen organization in AAT is unique from the organization in proximal scar tissue that was characterized by dense ribbons of linearly aligned thick collagen (Supplemental Figure 17b).

**Figure 6.**
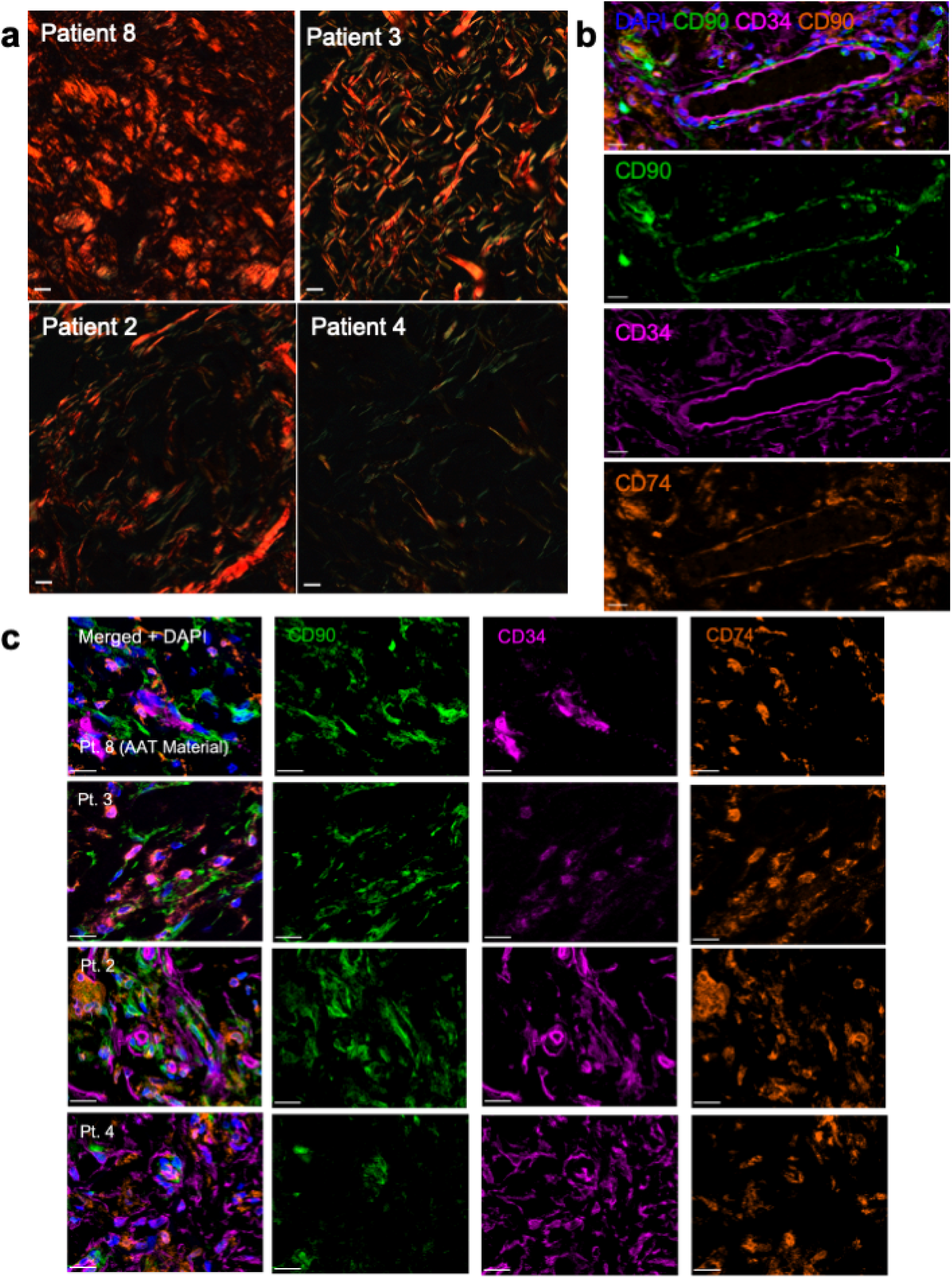
Collagen network remodeling analysis of AAT core-needle biopsies. **a.** Polarized light images of PSR staining of AAT material to investigate collagen network remodeling. Scale bar=10 µm **b.** Multiplex immunofluorescence staining of progenitor marker (CD34^+^), stromal marker (CD90^+^), and receptor for macrophage inhibitory factor (MIF) (CD74^+^) revealed pre-mature endothelial cells in AAT material important for angiogenesis. Scale bar=20 µm **c.** Multiplex immunofluorescence staining of progenitor marker (CD34^+^), stromal marker (CD90^+^), and receptor for macrophage inhibitory factor (MIF) (CD74^+^) to confirm presence of anti-fibrotic CD74^+^ ASCs (CD34^+^CD90^+^CD74^+^) in AAT Material. Scale bar=20 µm

Previous characterization of AAT demonstrated that there are soluble factors from AAT that attract adipose-derived stromal cells (ASCs) *in vitro*. Phase I AAT explants revealed CD34^+^ progenitor cell infiltration *in situ* [21]. To further examine differentiation potential in AAT in a clinically relevant cohort, immunofluorescence staining of CD34 marker was employed. Cell contact interactions were investigated with the additional staining of stromal cells CD29 and T cell marker CD3 (Supplemental Figure 18a, Supplemental Figure 19a). The proportion of biopsy by tissue type and total area analyzed were shown in Supplemental Figure 19b. CD34 is expressed in hematopoietic stem cells/progenitor cells and can also be expressed by endothelial cells. CD34 is enriched in pre-mature endothelial cells and fibroblast subtypes. CD34^+^ cells were present in all AAT biopsies and were abundant in Pt. 2 and Pt. 4 (Supplemental Figure 18b). CD34 expression in AAT biopsies demonstrates replicative capacity and differentiation potential. Cell-ECM interactions are key for stem cell fate determination. Other CD29^+^ stromal cells and CD3^+^ cells were present in AAT material. CD3^+^ T cells were found in close contact with CD34^+^ cells. CD3^+^ T cells also had contact with HLA-DR^+^ cells (Supplemental Figure 19c).

There are known protective roles of CD74 in injury and wound healing. MIF-CD74 signaling activates pro-survival and proliferative pathways resulting in protective and anti-fibrotic effects. Previous studies have demonstrated lipofilling or fat grafting can improve scar quality and clinical appearance. These desirable outcomes may be related to progenitor activity [30]. Cell-assisted lipotransfer with CD74^+^ adipose-derived stromal cells in a radiation-induced fibrosis skin preclinical model mediated antifibrotic effects [31]. A preclinical study with an acellular adipose tissue product demonstrated ECM-based products can also improve radiation-induced skin fibrosis (RIF) [32]. HA biomaterial-CD44 signaling also mediated survival pathway activation [33]. To evaluate if antifibrotic CD74^+^ adipose derived stromal cells (ASCs) were present in AAT, immunofluorescence staining was employed on biopsies with CD90, CD34, and CD74 markers. AAT material promotes progenitor activity and pre-mature endothelial cells important for angiogenesis (Fig. 6b). CD74^+^ ASCs were abundant throughout AAT regions of the biopsy samples (Fig. 6c).

### Discussion

Regenerative medicine technologies have the goal of functionally restoring tissues that have been damaged due to injury or disease; however, their successful progression to commercialization has been slow. Clinical translation and commercial ECM-based biomaterial indications vary widely [34]. Promising preclinical outcomes, mainly the induction of a pro-regenerative response characterized by type 2 immunity for ECM-based biomaterials for soft tissue injury applications, have not successfully been translated clinically for large volume soft tissue defects [35, 36]. Adipose-derived ECM-based biomaterials are good candidates for soft tissue reconstruction due to the abundance of fat tissue from cadaveric sources [37]. ECM from adipose is also appropriate because of its tissue-specific protein composition. ECM from adipose has ECM-specific structural and functional cues to support adipogenesis and angiogenesis. In this study we characterized the host response to permanently placed AAT, an off-the-shelf, adipose-derived biomaterial that can be injected in the clinic to reconstruct soft tissue defects. The first 10 patients completed the Phase II clinical trial, including one year of follow-up without any SAEs or unanticipated AEs associated with AAT treatment. AAT scored high for ‘naturalness’ in participant and investigator surveys—with results being more supportive of AAT use in the chest defects, this suggests that modest volumes of AAT may be better suited for restoring defects in the chest and larger volumes may be needed to restore abdominal defects. PRA lab testing showed that there was no increase in HLA antibodies at 4 weeks or 12 months post-injection for any patient, demonstrating that AAT does not cause a systemic immunologic response in concordance with Phase I findings. Histological examination of core-needle biopsies by a pathologist found that cellular and tissue response to AAT material increased with time, including degradation and remodeling the presence of lymphocytes and giant cells, and the absence of congestion, mineralization, or edema fluid accumulation, was noted.

There is a spectrum of diverging tissue-specific immune responses to implanted materials associated with either regeneration or fibrosis that are relevant to musculoskeletal pathologies such as soft tissue reconstruction [22, 38–40]. The role of the innate immune system in regulating tissue fibrosis, degeneration, and repair has been recognized. Recent studies have also highlighted the critical involvement of the adaptive immune system in these processes [41–43]. Studies investigating the reciprocal crosstalk between the immune response and stromal cell populations during tissue injury and homeostasis have identified immune cell phenotypes as predictors of tissue regeneration outcomes. There are immune-related mechanisms of action (MOA) for tissue products that impact repair outcomes. Biological and synthetic biomaterial implants are associated with a spectrum of T cell responses and drive related production of cytokines interleukin (IL)-4 and IL-17, respectively [22, 39]. In this study we investigated the innate and adaptive immune response. With specific high-plex immunofluorescence staining, we found that AAT can induce tertiary lymphoid structures (TLS). TLS are important for localized immune activity and its presence suggests a functional adaptive immune response. The etiology of TLS formation is not fully understood. TLS are heterogeneous in composition and phenotype and can contribute to either disease progression or resolution depending on clinical context. The presence of TLS in the context of cancer and infection is typically favorable while it confers a negative prognostic factor in autoimmune and chronic inflammatory diseases [44]. In cancer immunotherapy development, ECM-based biomaterials have been proposed to induce TLS as a class of immunotherapies or to be used in combination with other immunotherapies [45]. AAT supports mature B cells as evidenced by the SpatialDecon results from the Nanostring DSP gene expression data. B cells are not traditionally thought of in wound healing beyond humoral protection. B cells also have other roles that may impact the tissue microenvironment like cytokine secretion, antigen presentation, and immune cell activation. Based on the SpatialDecon cell type estimates, plasma cells were more abundant than naïve B or B cell memory in all tissue types. Mature B cells, like plasma cells, are likely to respond to biomaterials in a pro-healing manner. Preclinical studies investigating the divergent B cell responses to ECM-based biomaterial and synthetic materials found that ECM biomaterials support the generation of differentiated B cells [11]. Antibodies secreted by plasma cells are known to enhance wound healing in an aseptic skin wound model. The humoral response effectively supports phagocytosis by opsonization of damaged tissue [46]. In this study, differences found in cell composition on based on tissue type may be due to differences in functional sites and adhesion molecules in AAT material. Further work is needed to test functionality and suitability of AAT-induced TLS in other potential applications.

Nanostring GEOMx spatial transcriptional data revealed unique cell types and gene expression profiles based on tissue type in clinical biopsies. Macrophages and naïve CD4^+^ T cells were the most abundant cell types in AAT. Gene expression analysis found that *EGR1, FOS, JUN, JUNB, MCL1,* and *NR4A1* were significantly differentially upregulated in AAT compared to both fat and scar. Upregulated genes in AAT were related to adaptive immunity and macrophage activity including phagocytosis, pro-survival, and angiogenesis. Notably, *EGR1*, *MCL1*, and *NR4A1* have known roles in angiogenesis, anti-apoptotic processes, and pro-inflammatory regulation, respectively. *EGR-1* can induce multiple tissue repair processes [47]. These data demonstrate that AAT remains immunologically active in clinically relevant patients and supports tissue repair.

Macrophages play important roles throughout the wound healing process. Distinct macrophage phenotypes are necessary for angiogenesis and all phenotypes can support angiogenesis in different ways [27, 48]. Macrophage polarization IF staining demonstrated that AAT promotes multiple macrophage phenotypes. In this study, AAT primarily contained double negative nonpolarized macrophages. When polarized, AAT generally promoted more alternatively activated CD206^+^ macrophages compared to CD80^+^ macrophages. CD206^+^ macrophages have known roles in remodeling phases of wound healing. Comparable frequencies of double-positive macrophages were present when CD80^+^ macrophages were present in biopsies. These results were different than the macrophage polarization flow cytometry results from the Phase I trial where there were primarily CD206^+^ polarized macrophages in AAT explants [21]. This could be due to differences in time points; the Phase I study investigated explants up to 6 weeks *in situ* while the Phase II study evaluated core-needle biopsies up to 12 months after initial injections. In preclinical studies mixed macrophage phenotypes were found to be important to achieve vascularization of tissue engineering scaffolds [27, 49]. Aging-induced immunological changes and the nature of complex defects sites influence the spectrum of host responses to biomaterials, thus make it challenging to identify optimal patient populations and indications. A few senescent p16^+^ cells were present in some AAT biopsies. p16^+^ cell density was much higher in scar tissue compared to AAT. Further work in understanding the role of SASP factors and senescence in wound healing is needed.

The local microenvironment and extracellular matrix have important roles in cell adhesion, cell signaling, proliferation, and cell fate determination. The collagen organization in AAT biopsies investigated with PSR staining showed distinct collagen organization compared to proximal scar tissue. AAT regions were characterized by thin immature collagens randomly organized and not densely packed while proximal scar had densely packed and linearly organized thick collagen. Based on the significant upregulation of *CD74* in AAT compared in fat in the Nanostring DSP data and the lack of dense collagen organization in AAT regions we hypothesized that AAT may promote CD74^+^ ASCs. CD74 has known roles in mediating repair in different organs and tissue during injury. IF staining confirmed the presence of CD74^+^ ASCs. CD74^+^ ASCs have been shown to have increased expression of antifibrotic growth factors (FGF-2, hepatocyte growth factor) [31]. This study demonstrated that AAT alone could promote CD74^+^ ASCs. ASCs in stromal vascular fraction (SVF) of lipoaspirate are associated with conferring the main therapeutic action of fat grafting [50, 51]. Here, we show that AAT supports angiogenesis and CD34^+^ progenitor cell recruitment without being used in conjunction with growth factors or autologous progenitor cells. ASCs are important for material integration and tissue repair.

The limitations of the study are acknowledged. The study is not complete, additional patients will be recruited. The trial had a limited number of patients and thus findings from the study may not apply broadly. The use of ultrasound-guidance for biopsy acquisition aimed to mitigate sampling error. The defects are of modest size and biopsy samples only capture a small fraction of the defect site at a snapshot in time. Limited assays were conducted due to the small size of the biopsies. Explants are typically used to study complete host response; yet this was not possible because AAT was permanently placed for soft tissue reconstruction. However, biopsies are routinely used to confirm prognosis and diagnosis and thus can elucidate valuable information. Importantly, the study found biological differences between AAT and proximal tissues and valuable insights in a clinically relevant population. The study validated preclinical and Phase I results that showed that AAT promotes angiogenesis and progenitor activity. In some biopsy acquisitions, there was inadequate total tissue capture or inadequate capture of regions with AAT material. It was not possible to biopsy all patients due to prior medical history of one patient. The study protocol only indicated one biopsy per patient and thus was not able to follow the remodeling process of each patient with repeated biopsies. There were numerous biological variables including differences in AAT volumes administered to each patient, anatomical location, and nature of the soft tissue defect, and radiation history status. There were also gender and age differences among patients that could impact the tissue repair outcomes. Nonetheless, the study found differentially expressed genes by tissue type. Overcorrection is a common practice in fat grafting to compensate for anticipated fat resorption where surgeons fill the volume of the defect beyond its size. There were challenges in finding patients with exact size defect for each volume cohort needed in the study. Thus, defects could not always be filled because of specific volumes based on cohort size in the study design. This could have implications related to survey results.

This study provides insight into host response to AAT in a clinically relevant population and supports product development. AAT maintains its regenerative capacity to promote endogenous immune and progenitor cell recruitment that ultimately support angiogenesis and remodeling at a soft tissue injury site. This work has important implications for current clinical practice and will impact reconstruction methods and treatment options in multiple clinical fields. The host response to AAT in patients with complex defects due to histories of radiation and cancer is unknown. Different anatomical locations can result in different tissue-specific responses. This study opens the door to continue to evaluate the use of AAT in patients with complex soft tissue defects like those with previous history of radiation. Future studies should investigate additional defect sites and further profiling with techniques such as scRNA seq to identify molecularly distinct populations. Patients that have scar tissue or radiation-induced changes to defect site can have limited reconstructive options. Additionally, there are no current treatment options for smaller defects such as lumpectomy that are disfiguring. The study showed that AAT implantation supports pro-regenerative and anti-fibrotic response regardless of surrounding tissue. Further success in the translation of biomaterials will require understanding the spatial and temporal host response and the role of specific ECM components. There has been limited clinical impact using regenerative medicine strategies in part due to lack of understanding of their mechanism of action and their complex composition. This study will impact patient care and inform future development of next generation ECM-based biomaterials.

### Methods

#### Contact for Reagent and Resource Sharing

Further information and requests for reagents or more information may be directed to and will be fulfilled by the corresponding author Jennifer H. Elisseeff.

#### Ethics statement

A Phase II, dose-escalation, open-label study (NCT03544632) evaluating the safety and efficacy of permanently placed acellular adipose tissue (AAT) in human subjects with modest soft tissue defects of the trunk was conducted at Johns Hopkins University with IRB and DOD OHRO (formerly known as HRPO) approvals. Human core needle biopsy samples (14-gauge hollow needle) were obtained with approval from the Johns Hopkins Medicine (Baltimore, MD) IRB (Study ID# IRB00155003) and the OHRO Log No. E01422.1a.

#### Manufacturing clinical AAT

AAT was manufactured from aseptically harvested cadaveric adipose tissue under aseptic processing with adherence to 21 CFR 210 & 211, Current Good Manufacturing for Finished Pharmaceuticals, and ICH Q7, Good Manufacturing for Active Pharmaceutical Ingredients. AAT is terminally sterilized with gamma irradiation. Donor tissue was screened for suitability and acquired from an organ procurement organization.

#### Subject Recruitment and Enrollment

2,400 patients were screened for eligibility. Ten patients were enrolled based on inclusion and exclusion criteria (Supplemental Table 9). The first cohort of patients had soft tissue defects that were approximately 4 ccs (n=5), the second cohort had soft tissue defects that were approximately 10 ccs (n=5). Reinjections (up to 40 cc total injection volume) were optional 3 months after initial injection. All subjects signed informed consent forms (ICF) that outlined potential risks to participate in the JHM IRB– and OHRO-approved study prior to engaging in any study-related activities.

#### Clinical Trial Design

The clinical study was a 12-month, prospective, Phase II, dose escalation study assessing the safety and efficacy of permanently placed AAT intended for repair of modest (approx. 5-30cc) soft tissue defects of the chest or trunk. It included three maximum injection volume groups (5 cc, 10 cc, and 20 cc). For a maximum injection volume not to exceed 40 cc. CONSORT (Consolidated Standards of Reporting Trials) Flow Diagram is shown in Supplemental Figure 1 to outline patient enrollment and follow-up visits. Ten patients (7 females, 3 males) aged 20-65 (mean: 49) years enrolled in and completed the dose-escalation, open-label study. Inclusion and exclusion criteria are listed in Supplemental Table 12.

#### Study Objectives

The primary objective was to assess the safety and efficacy of acellular adipose tissue (AAT) to restore volume in soft tissue defects of the trunk. Secondary objectives were to assess histopathology of implant core needle biopsies up to 12 months post initial injection and tolerability of the material.

#### Administration of AAT

AAT injections were done under local anesthesia and sterile conditions using a blunt needle. AAT was injected in soft tissue defects of the chest or

trunk using a “fanning” technique like that typically used in fat grafting and filler injection procedures. The “fanning” technique integrates AAT with the surrounding tissue.

#### Clinical Assessment at Follow-up Visits

Patients had 14 total follow-up visits during the study comprised of eight (n=8) in-person visits and six (n=6) safety calls. In-person follow-up visits included vitals, clinical labs, assessments, imaging, surveys, review of concomitant medications, and recording of any unanticipated or adverse events. Safety calls consisted of surveys, review of concomitant medications, and recording of any unanticipated or adverse events. Core-needle biopsies were acquired (one per patient, as clinically appropriate) at one in-person follow up visit per patient as outlined in the study protocol. Longitudinal 2D clinical photographs may be made available upon reasonable request to the corresponding author, subject to appropriate ethical and institutional approvals.

#### Serious Adverse Events/Adverse Events

Serious adverse events (SAEs) and adverse events (AEs) were graded based on the Common Terminology Criteria for Adverse Events v4.0 (CTCAE) and reported during follow-ups. All AEs were captured on the appropriate study-specific care report forms (CRFs). AEs were graded from 1 to 5 (1 = mild to 5 = death related to the AE), and relationship assignment was determined to be one of the following per the clinical investigator and confirmed by the medical monitor: ‘not related’, ‘probably not related’, ‘possibly related’, ‘probably related’, or ‘definitely related’ to the AAT injection.

#### Ultrasound-guided Biopsy Acquisition

Ultrasound imaging was performed using a Esoate MyLab50 XVision system to facilitate identification of AAT. Core-needle biopsies were performed under local anesthesia. 14-gauge Max-Core disposable core-needle biopsy (BD) devices were used to obtain biopsies. Samples were fixed in 10% formalin solution overnight at 4 °C.

#### Safety and Tolerability

The primary objective of safety was determined by incidence and rate of adverse/unanticipated events. A secondary endpoint of assessing tolerability was determined by patient and physician surveys and panel reactive antibody (PRA) testing. PRA testing was performed at baseline prior to AAT injections, and 4 weeks and 12 months post-injections for each study participant to evaluate a change in IgG HLA.

#### Histopathology

Core-needle biopsies (one per patient, as clinically appropriate) were conducted at different regular time points up to 12 months after the initial injection as outlined in the study protocol. All samples were fixed in 10% formalin, processed, embedded in paraffin, and stained using Hematoxylin & Eosin (H&E) or Picro Sirius red staining (PSR) (Abcam, ab150681) according to standardized procedures. Whole slide H&E samples were imaged with Olympus VS200 Slide Scanner at 20x magnification. Regions were annotated by tissue type in HALO® (RRID:SCR_018350) Image Analysis Platform Version v3.4 (Indica Labs, Inc.) and the Multiplex IHC Module was used to calculate cell density; the total cell number was divided by the total area analyzed for each annotated region. An unblinded pathologist used a histopathological examination scoring system to score all H&E-stained samples derived from core needle biopsies.

#### Multiplex Immunofluorescence Staining

Samples were fixed and paraffinized using standard protocols. Immunofluorescence staining was employed on deparaffinized sections. Heat-mediated antigen retrieval were performed in appropriate citrate buffers (AR6 or AR9) (PerkinElmer) for 15 minutes in a vegetable steamer. Endogenous peroxidases were blocked using 3% H_2_O_2_ for 15 minutes. Slides were blocked with 10% bovine serum albumin (BSA) 0.05% Tween-20 in TBS-T (Sigma) for 30 minutes, then incubated with appropriate primary antibodies (Abcam) at optimized concentrations or appropriate isotype control at the same concentration in 10% BSA 0.05% Tween-20 in TBS-T ranging from 30 minutes at RT to overnight at 4°C. Appropriate HRP Polymer Detection systems were used according to manufacturer’s protocol to detect the antibodies of interest (BIOCARE MEDICAL, M3R531H or RH531H). Tyramide signal amplification (TSA) visualization was performed using appropriate Opal fluorophore (Opal 650, Opal 570, or Opal 520) (PerkinElmer) according to manufacturer’s protocol. Sections were rinsed with water then TBS-T, counterstained with DAPI for 5 minutes, rinsed, then mounted using DAKO mounting medium (Agilent), cover slipped, and subsequently imaged. A list of primary antibodies can be found in Supplemental Table 13.

#### Phenocycler Tissue Staining and Fixation

Complete methods for PhenoCycler formerly known as Co-detection by indexing (CODEX) experiments can be found in Black et al. [52]. Briefly, FFPE sampled were sectioned at 5-10 µm on poly-L-lysine coated coverslips. The samples were deparaffinized and antigen retrieval was performed. Samples were subsequently stained with an oligonucleotide-conjugated antibody panel. Samples were then washed and re-fixed prior to imaging. A PhenoCycler reporter plate was prepared with unique fluorescently labelled oligonucleotides complementary to each antibody in the panel. Samples were iteratively imaged at single cell resolution as fluorescently labelled oligonucleotides were added to the sample to detect the binding events with the corresponding antibody (up to three antibodies with different fluorophore channel and nuclear stain per cycle), then chemically stripped, washed, hybridized, and re-imaged for all antibodies in the panel. The panel used can be found in Supplemental Table 13. QuPath version 0.3.2 (RRID:SCR_018257) and (Fiji Is Just) ImageJ 2.3.0 (RRID:SCR_002285) were used for image processing.

#### Multispectral Image Acquisition and Opal library creation

Multispectral 20X magnification images were acquired using the Vectra Polaris Automated Quantitative Pathology Imaging System using the Multispectral slide scan (MOTiF) method as suggested by the manufacturer (Akoya Biosciences). Whole slide scans were selected for inForm batch analysis in Phenochart 1.1.0 (RRID:SCR_019156). For each panel an Opal library was created using appropriate single-fluorophore library slides and a slide with no stain for autofluorescence was imaged and high-power fields were selected in Phenochart 1.1.0 or synthetic libraries were used. Once created, the Opal library was used for multispectral unmixing of samples in inForm 2.5.1 (RRID:SCR_019155) to isolate emission from each fluorophore.

#### Quantitative Image Analysis

We performed cell phenotyping and specific cell type density quantification of Vectra opal multiplex immunofluorescence imaging using HALO® (RRID:SCR_018350) Image Analysis Platform Version v3.4 (Indica Labs, Inc.). Each biopsy was annotated by tissue type (AAT, scar, or fat tissue). For H&E cell density quantification, single-cell segmentation and cell-type quantification was performed using the Multiplex IHC module v2.3.4. For all quantitative analysis for immunofluorescence staining, HighPlex FL module v3.2.1 was used. Cell density by desired cell phenotype was calculated by dividing the number of total desired cell type by total area analyzed. Regions of biopsies were excluded from analysis if folded tissue or other irregularities were present. Representative images were taken at 20× magnification on an Axio Observer.Z1 (Zeiss) microscope with Axiocam 305 color imaging device and Apotome.

#### Picrosirius Red Imaging and Image Analysis

20× magnification polarized light images were acquired using ZEISS Axio Imager 2 Pol microscope. Tile images were acquired for the entire biopsy samples. Representative polarized light images were taken at 20× magnification on an Axio Observer.Z1 (Zeiss) microscope with Axiocam 305 color imaging device.

#### Spatial RNA Profiling in situ Hybridization and Sequencing

Comprehensive methods for GEOMx Digital Spatial Profiler (DSP) (RRID:SCR_021660) RNA assays can be found in Merritt et al. [53]. Briefly, FFPE samples were re-embedded and freshly sectioned at 4 µm on Leica Bond Plus slides. Sections were dried overnight at 37°C. Sections were stored in a vacuum-sealed container at 4°C until the assay was performed. Immunofluorescence staining CD45, CD31, CD68, and DNA (CYTO13) and RNA detection probes with UV-photocleavable oligonucleotide containing a unique molecular identifier (UMI) using standard immunohistochemistry protocols were employed using GEOMx RNA Slide Prep FFPE PCLN. The tissue section was imaged to generate a whole tissue image at single cell resolution. Regions of interest (ROI) were identified based on morphology staining and histologic examination from serial H&E staining. Each ROI was segmented into two regions: CD31^+^ and CD31^-^ regions. Samples were washed then loaded on the GEOMx Digital Spatial Profiler (DSP). 192 distinct circular ROIs were sequentially collected by photocleaving the oligonucleotides from the profiling and indexed to the ROI on the tissue. Oligonucleotides were hybridized overnight at 37°C to code set tags, 1,800+ gene probes using the Cancer Transcriptome Atlas (CTA) (Nanostring) and quantified using Next Generation Sequencing (NGS) (Illumina) for high profiling gene expression data. The assay was performed by the Cytometry Facility at the UPMC Hillman Cancer Center.

#### Nanostring GEOMx DSP Data Processing and analysis

Data analysis was performed in R. Visualizations were made using Prism 9, and R packages Complex Heatmap and ggplot2. Data were processed using the technical and biological probe quality control parameters according to the manufacturer’s protocols. Probes were filtered from the data if the ratio of the geomean probe in all segments) by the geomean probes within target) is less than or equal to 0.1 or if probes fail Grubbs outlier test where probes should not be an outlier in greater than or equal to 20% of the segments.

The limit of quantification (LOQ) filtration, a confidence threshold defined as the negative probe geomean multiplied by the geometric standard deviation of negative probes two standard deviations above geomean of the negative probes, were employed for genes and segments. Data were normalized using the variance stabilizing transformation (vst) function from DESeq2 [54]. Differential gene expression was performed using a linear mixed model as implemented by Limma [55]. For all comparisons, patient identity was set as a random effect, and significant genes were identified with significance thresholds of p_adj_ < 0.05 and 0.58 ≤ LFC or ≤ –0.58. Spatial cell deconvolution of the vst normalized GEOMx DSP data was performed using the SpatialDecon R package [13] with the ImmuneTumor_safeTME normalized cell matrix from Nanostring (<u>Nanostring-Biostats Github</u>). PPI networks of differentially expressed genes made up of high confidence (0.7) both functional and physical protein associations were created using STRING v. 11.5 (RRID:SCR_005223) and visualized using Cytoscape 3.9.0; disconnected nodes in the network were hidden [17]. Enrichment detection was employed using an automated pathway enrichment analysis from STRING. Overrepresentation tests using multiple functional pathway classification frameworks were used: Gene ontology annotations, UniProt Keywords, and REACTOME Pathways [56–58]. False discovery rate (FDR) values from the functional enrichment analysis of PPI were corrected for multiple testing within each category using the Benjamini–Hochberg procedure [59]. The R script used for analysis is included in the supplemental files.

#### Statistical Analysis

Statistical analyses were performed using GraphPad Prism 9.0.0 and R. Statistical analyses of the SpatialDecon results were performed using the ordinary one-way ANOVA with Tukey’s multiple comparisons test. All statistical tests, alpha=0.05.

#### Materials Availability

There are restrictions to the availability of acellular adipose tissue (AAT) product due to its status of being clinically evaluated and not yet available for commercial use.

***Data and Code Availability***. All relevant data are included in the manuscript or in the supplementary material.

#### Author Contributions

Conceptualization, J.H.E., D.S.C., P.B. C.M.C.; Formal analysis, A.N.P., A.T.F.B., E.J.F., J.C.S.; Investigation, A.N.P., J.A.G.; Resources, A.E.A.; Writing—Original Draft, A.N.P; Writing—Reviewing and Editing, all authors; Supervision, J.H.E., D.S.C.; Project Administration C.M.C., P.A., J.P.; Funding Acquisition, J.H.E.

### Declaration of Interests

J.H.E. holds equity in Aegeria Soft Tissue, a company that has licensed JHU intellectual property on an acellular adipose tissue (AAT) product for soft tissue reconstruction. The conflict is being managed by the Johns Hopkins Office of Policy Coordination. J.H.E. is an inventor on patents related to the AAT technology. P.B. is the Chief Medical Officer of Aegeria Soft Tissue and holds equity. At the time of study, the conflict was managed by the Johns Hopkins Office of Policy Coordination. All other authors declare that they have no competing conflict of interest.

## Data Availability

All data produced in the present study are available upon reasonable request to the authors

## Acknowledgements

This research was supported by the Armed Forces Institute for Regenerative Medicine (AFRIM II) and the Department of Defense (DoD) Clinical Trial Award (CTA). A.N.P. was supported by NSF-GRFP award #DGE-1746891 and J.A.G. was supported by a T32 award #T32DC000027-29 at the time of study. The Immune Microenvironment Lab core at Johns Hopkins School of Medicine aided in Vectra Polaris imaging and insight with image processing. E.J.F. was supported by the Cancer center core grant, P30CA006973, at the time of study. J.C.S. was supported by Dermatology Foundation Career Development Award, Johns Hopkins Clinician Scientist Award, and NIH P41EB028239 at the time of study. This project used the Hillman Cancer Center Cytometry Facility that is supported in part by award P30CA047904. Figures were created with BioRender.com

---

**Supplemental Figure 1.**
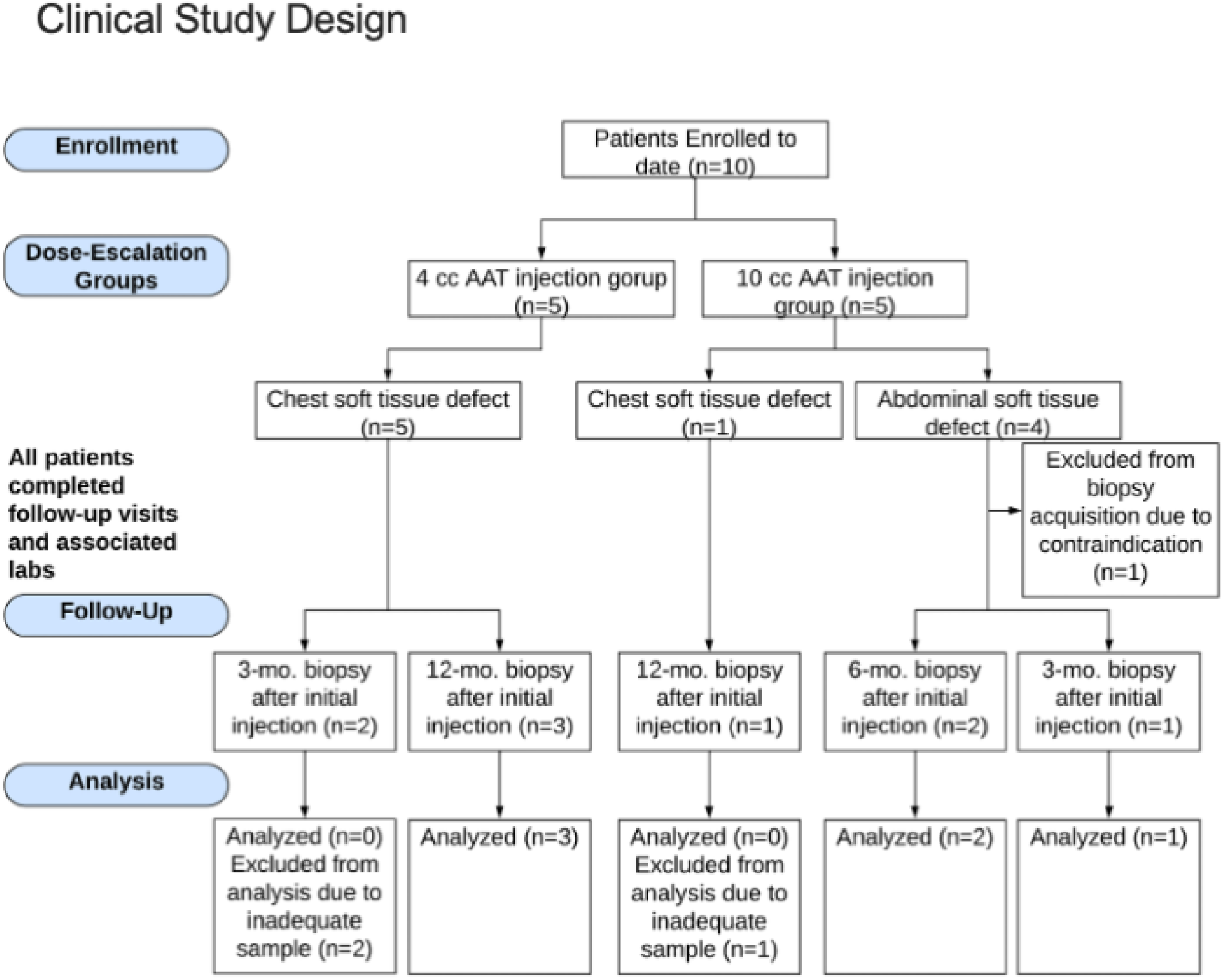
Clinical Study Design using CONSORT (Consolidated Standards of Reporting Trials) Flow Diagram.

**Supplemental Figure 2.**
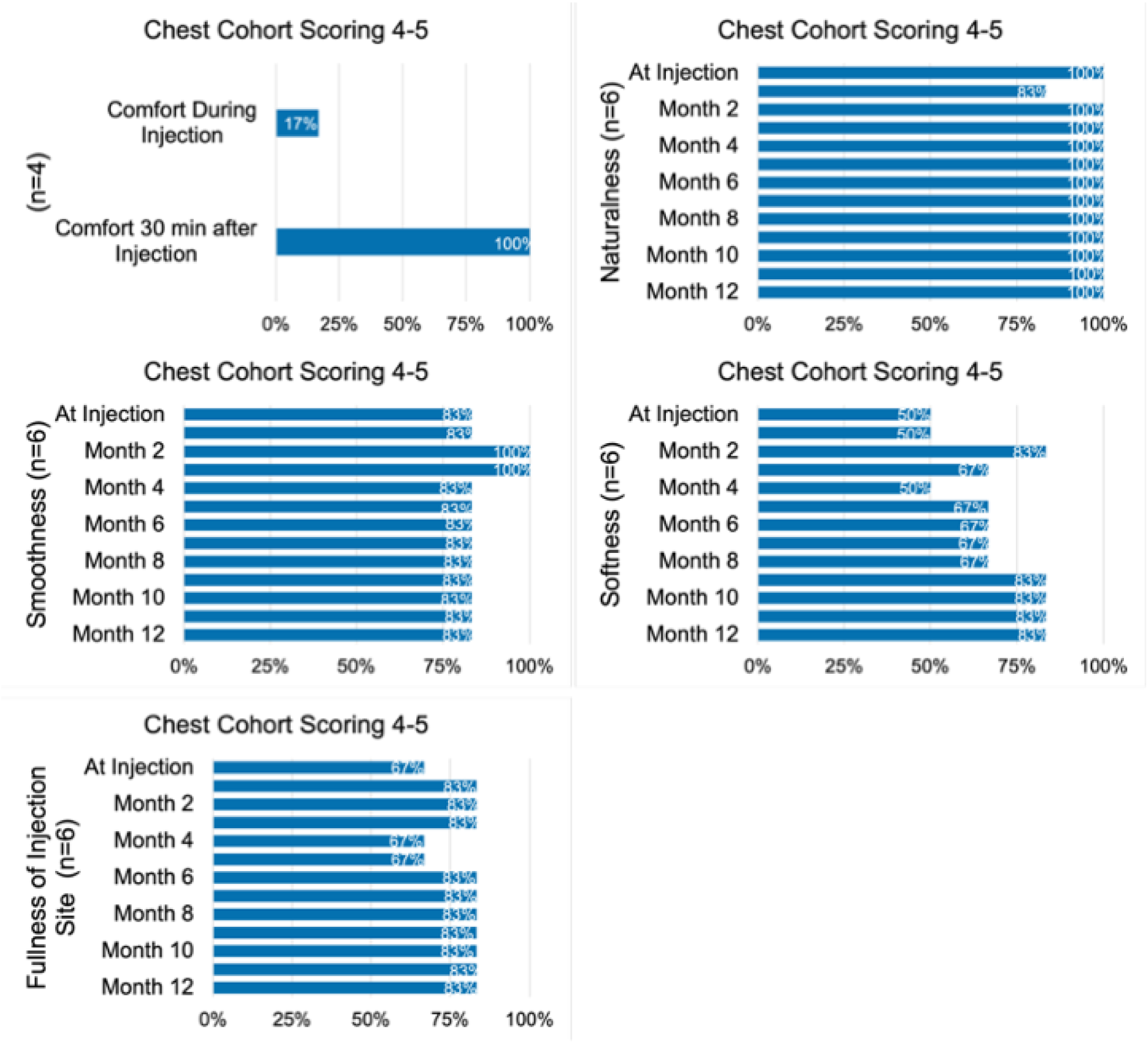
Participant surveys. Participant comfort and appearance survey results for the Chest cohort obtained at injection and follow up visits. Percentage of patients rating scores 4 or 5 on the survey indicating satisfaction.

**Supplemental Figure 3.**
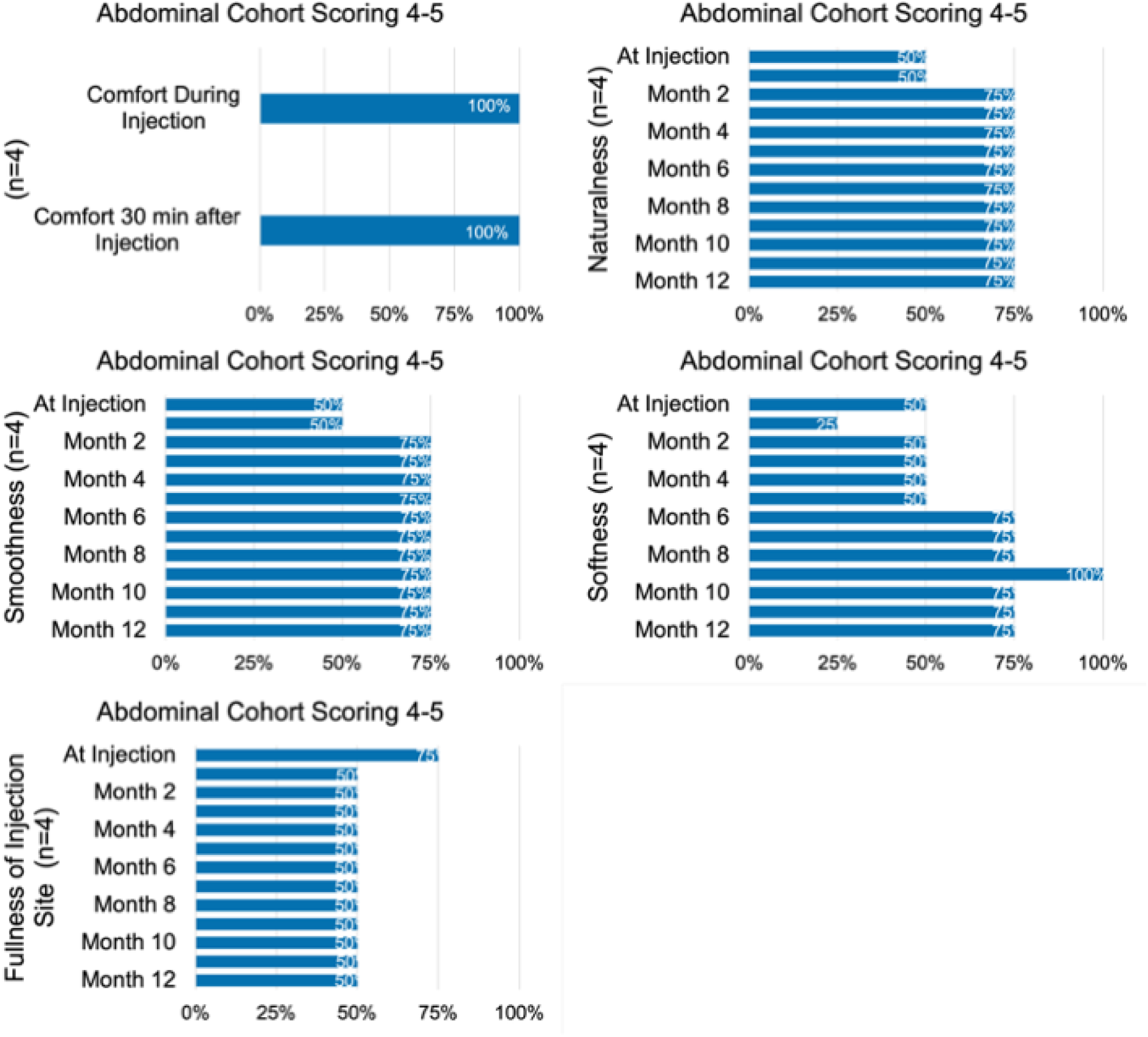
Participant surveys. Participant comfort and appearance survey results for the Abdominal cohort obtained at injection and follow up visits. Percentage of patients rating scores 4 or 5 on the survey indicating satisfaction.

**Supplemental Figure 4.**
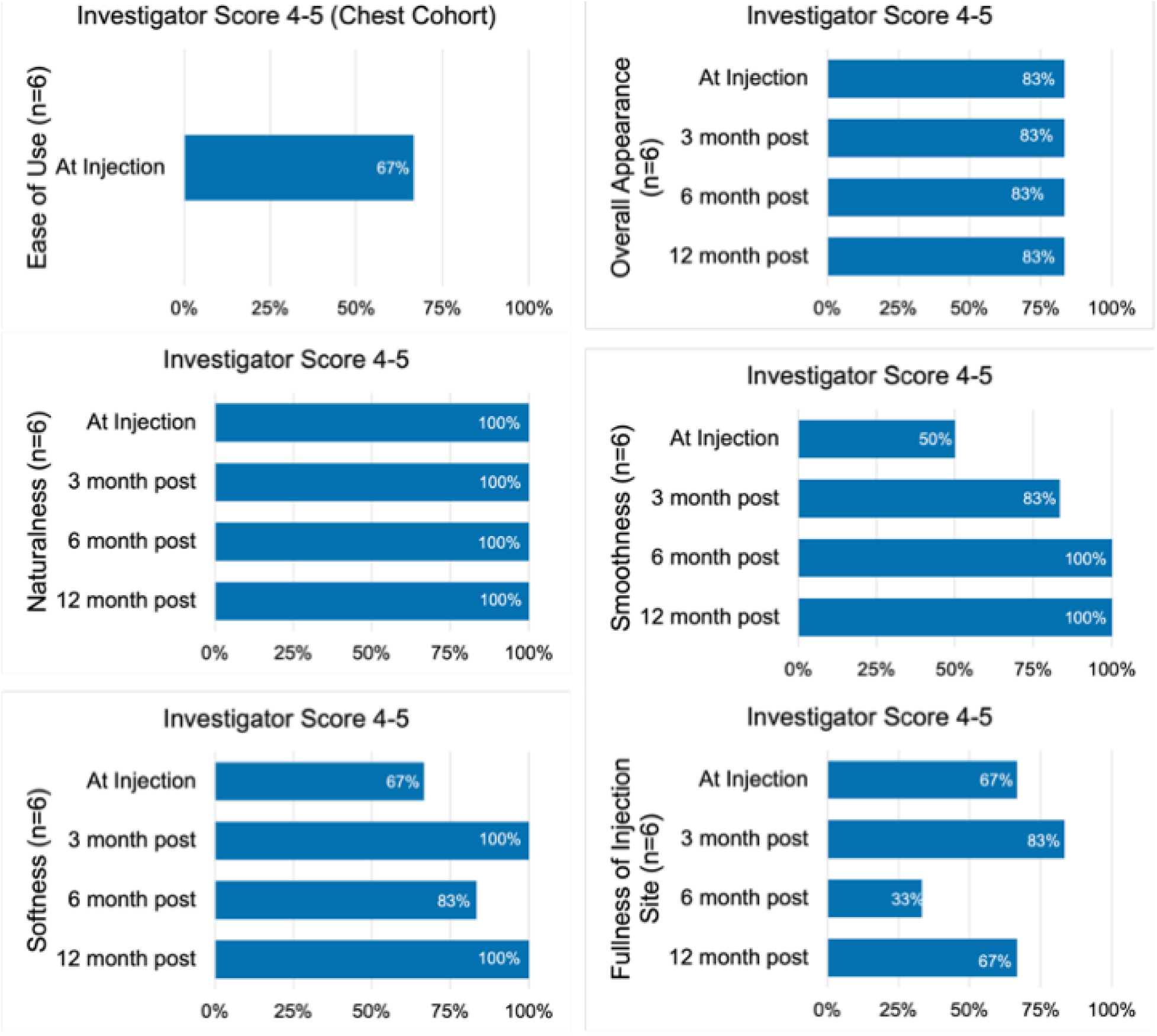
Investigator surveys. Investigator ease of use and appearance survey results corresponding to the Chest cohort obtained at injection and follow up visits. Percentage of scores 4 or 5 on the survey indicating satisfaction by the investigator.

**Supplemental Figure 5.**
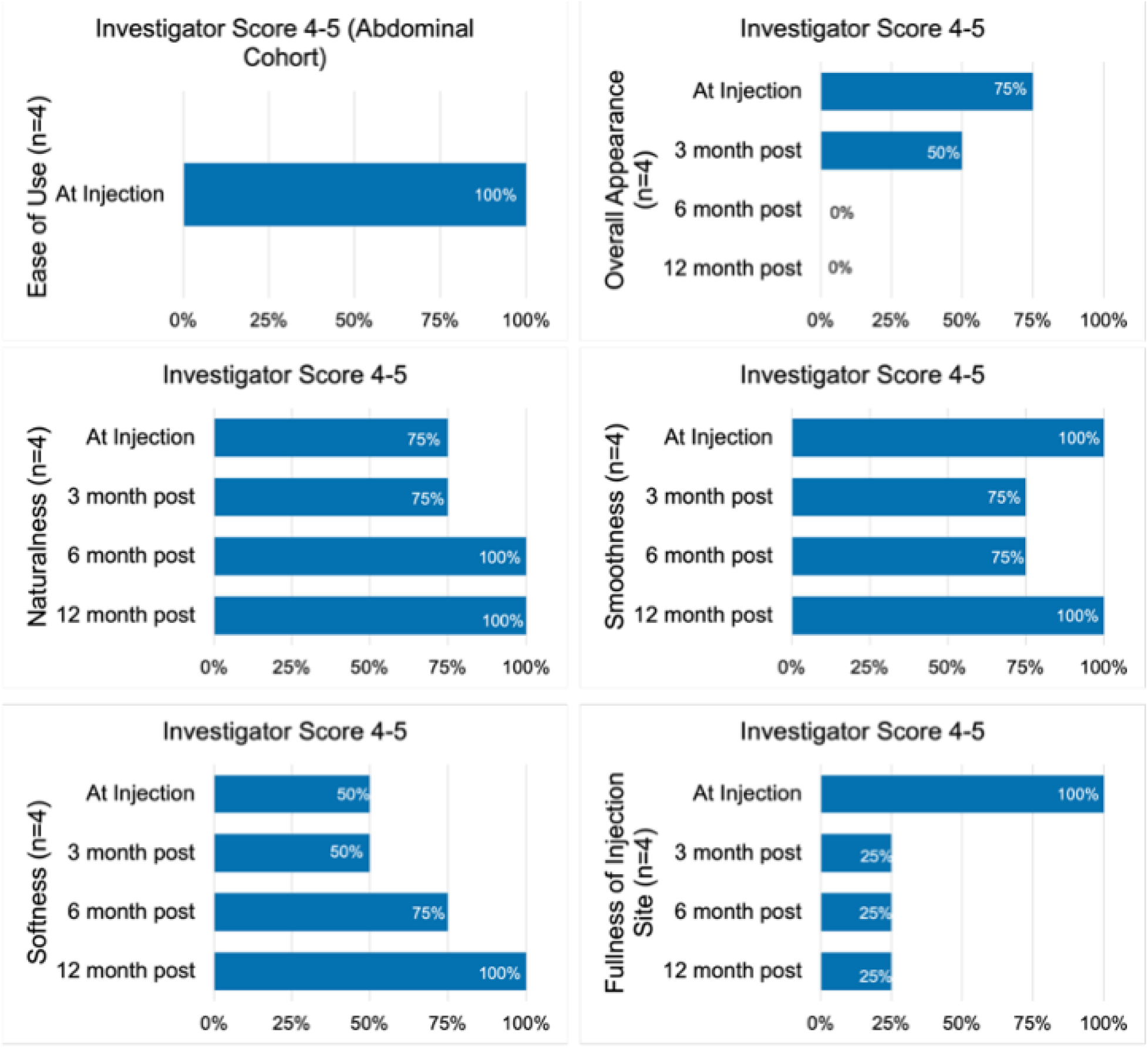
Investigator surveys. Investigator ease of use and appearance survey results corresponding to the Abdominal cohort obtained at injection and follow up visits. Percentage of scores 4 or 5 on the survey indicating satisfaction by the investigator.

**Supplemental Figure 6.**
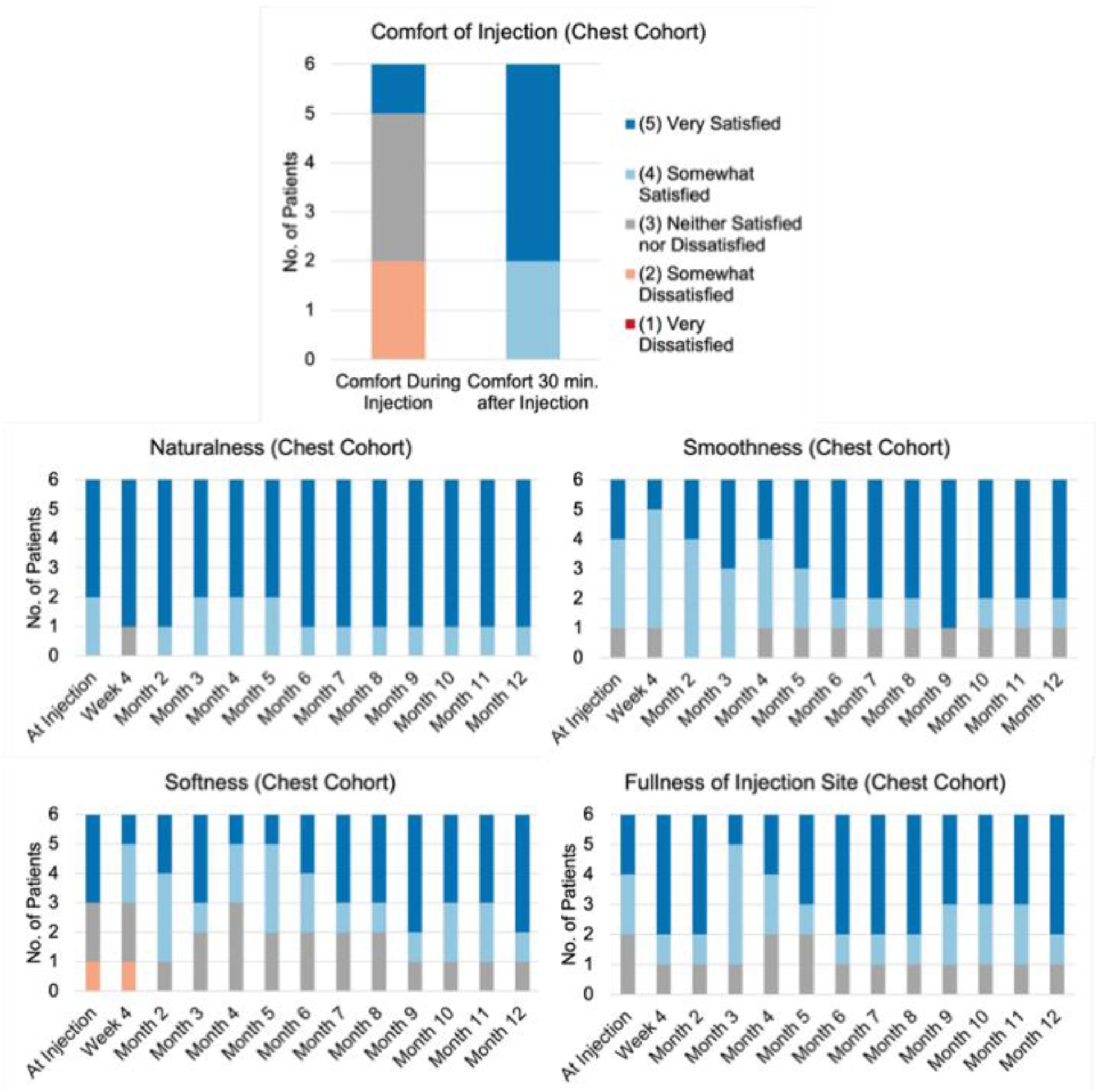
Participant surveys. Participant comfort and appearance survey results for the Chest cohort obtained at injection and follow up visits.

**Supplemental Figure 7.**
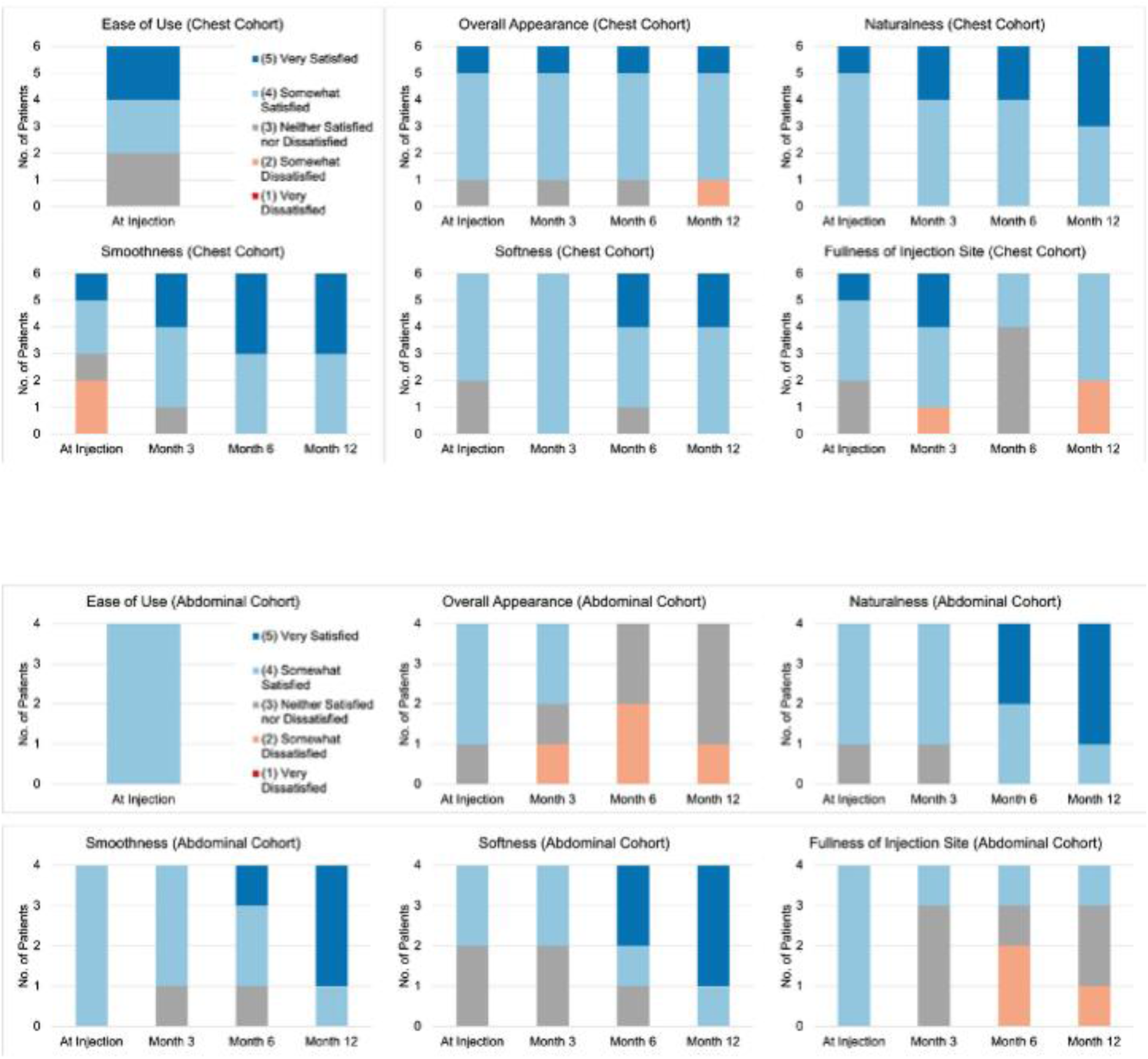
Investigator surveys. Investigator ease of use and appearance survey results corresponding to the Chest and abdominal cohort obtained at injection and follow up visits.

**Supplemental Figure 8.**
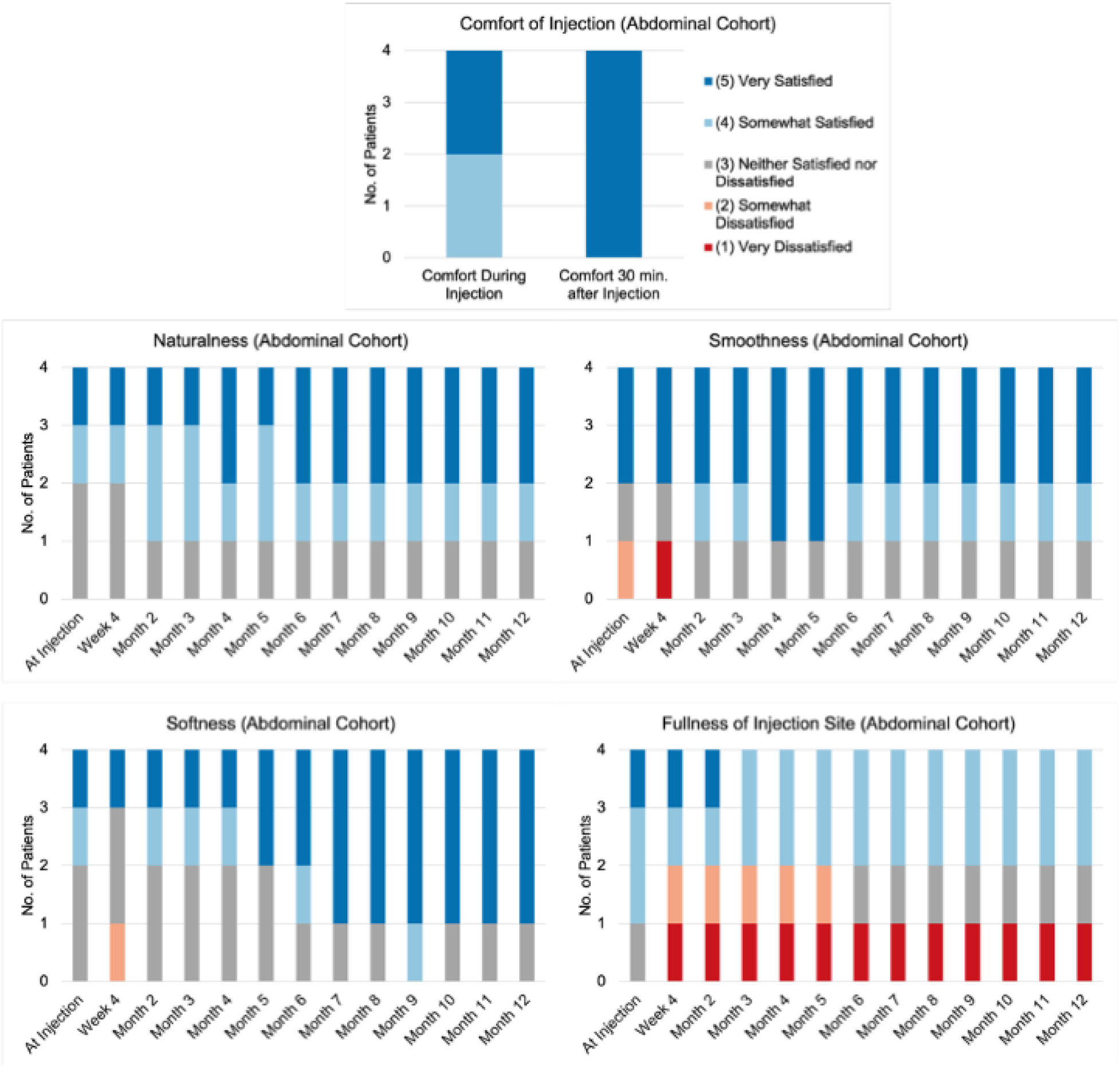
Participant surveys. Participant comfort and appearance survey results for the abdominal cohort obtained at injection and follow up visits.

**Supplemental Figure 9.**
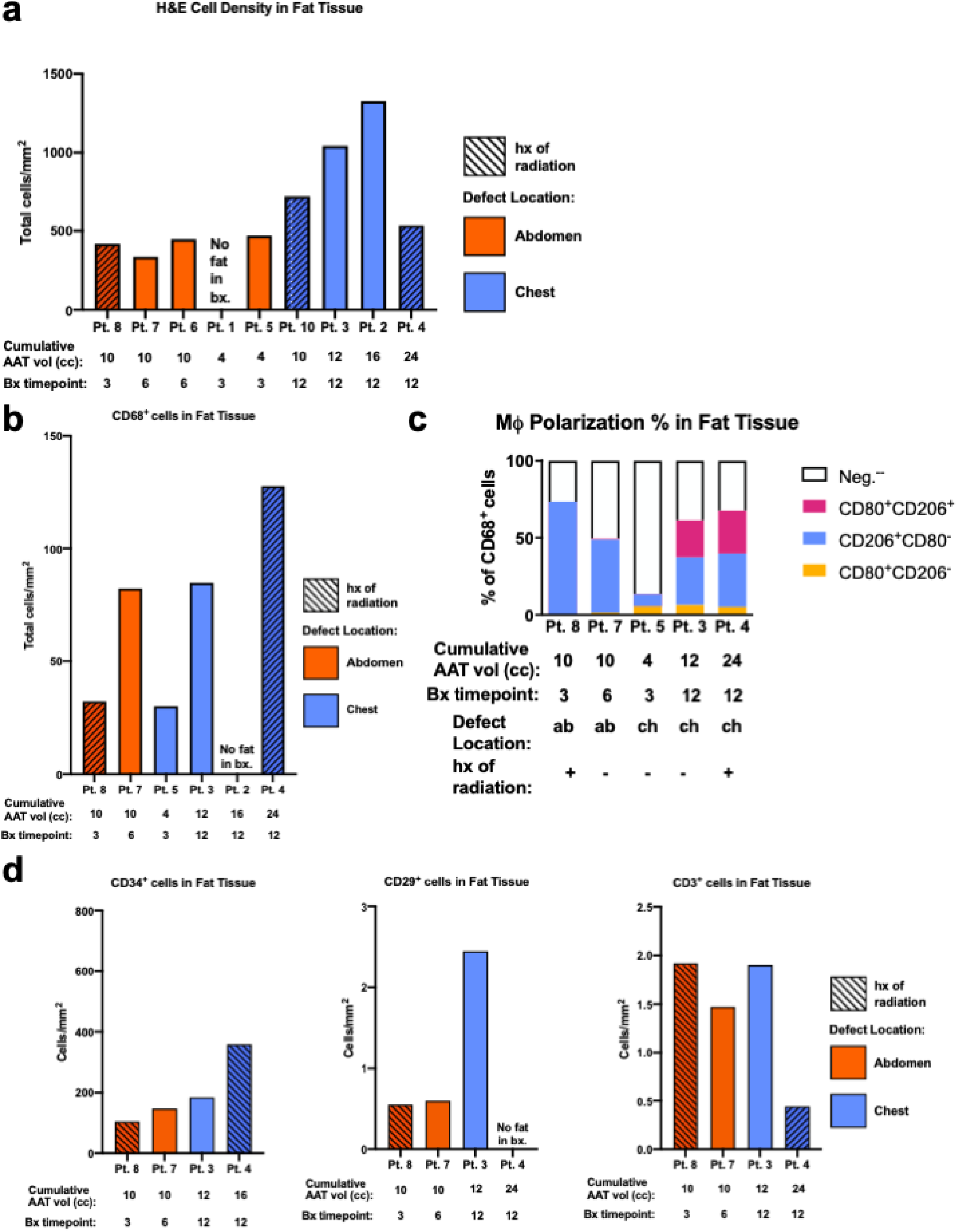
Fat Tissue Data. **a**. H&E Cell density quantification in fat tissue quantified using HALO **b.** CD68^+^ cell density in fat tissue quantified by IF staining using HALO **c.** Macrophage polarization in fat tissue quantified by IF staining using HALO. Populations of nonpolarized (CD68^+^CD80^-^CD206^-^ cells), classically-activated (CD68^+^CD80^+^CD206^-^ cells), (CD68^+^CD206^+^CD80^-^cells), and double-polarized (CD68^+^CD80^+^CD206^+^ cells) represented as a percentage of CD68^+^ cells **d.** CD34^+^ cell density, CD29^+^ cell density, and CD3^+^ cell density in fat tissue quantified by IF staining using HALO

**Supplemental Figure 10.**
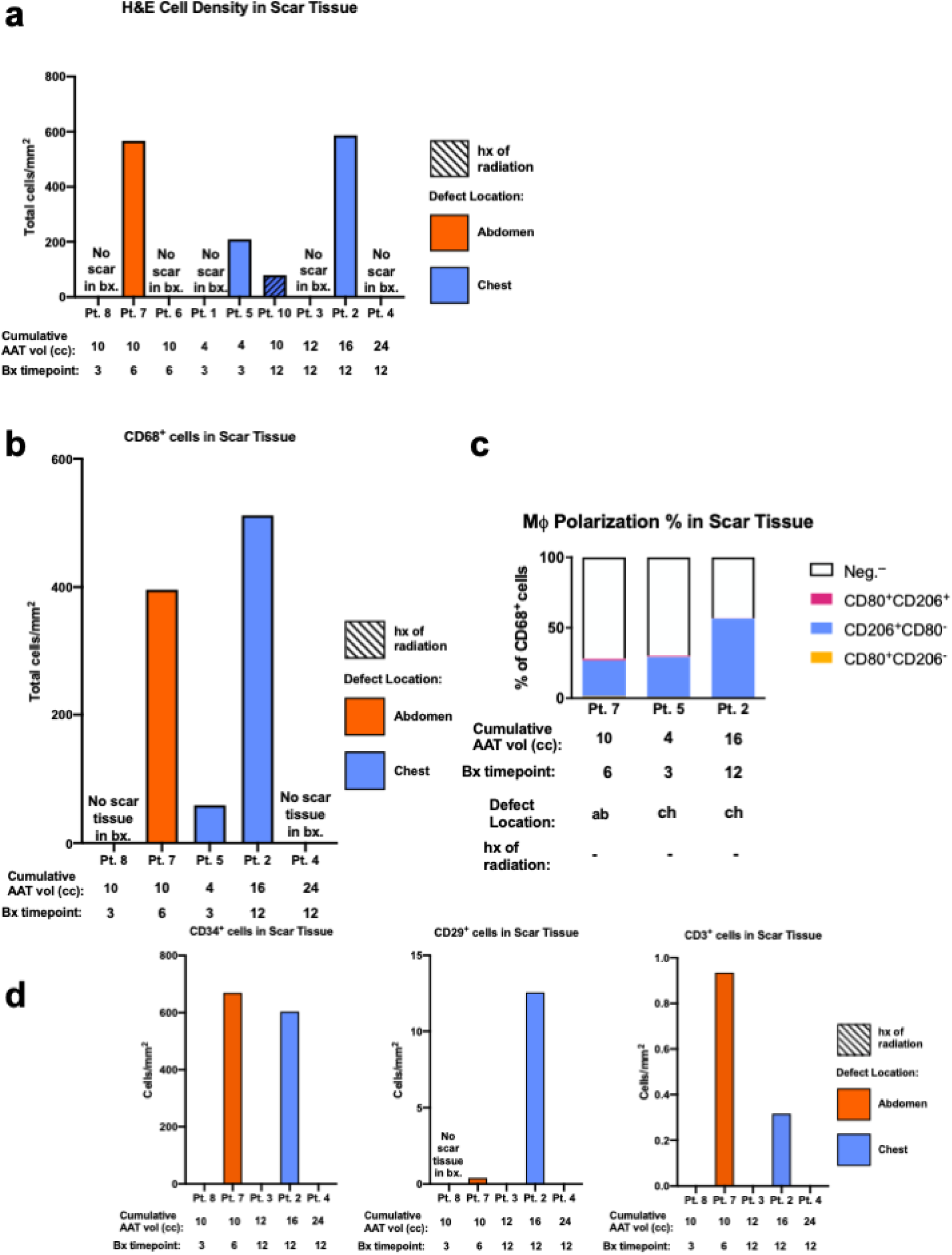
Scar Tissue Data. **a.** H&E Cell density quantification in scar tissue quantified using HALO **b.** CD68^+^ cell density in scar tissue quantified by IF staining using HALO **c.** Macrophage polarization in scar tissue quantified by IF staining using HALO. Populations of nonpolarized (CD68^+^CD80^-^CD206^-^ cells), classically-activated (CD68^+^CD80^+^CD206^-^ cells), (CD68^+^CD206^+^CD80^-^ cells), and double-polarized (CD68^+^CD80^+^CD206^+^ cells) represented as a percentage of CD68^+^ cells **d.** CD34^+^ cell density, CD29^+^ cell density, and CD3^+^ cell density in scar tissue quantified by IF staining using HALO

**Supplemental Figure 11.**
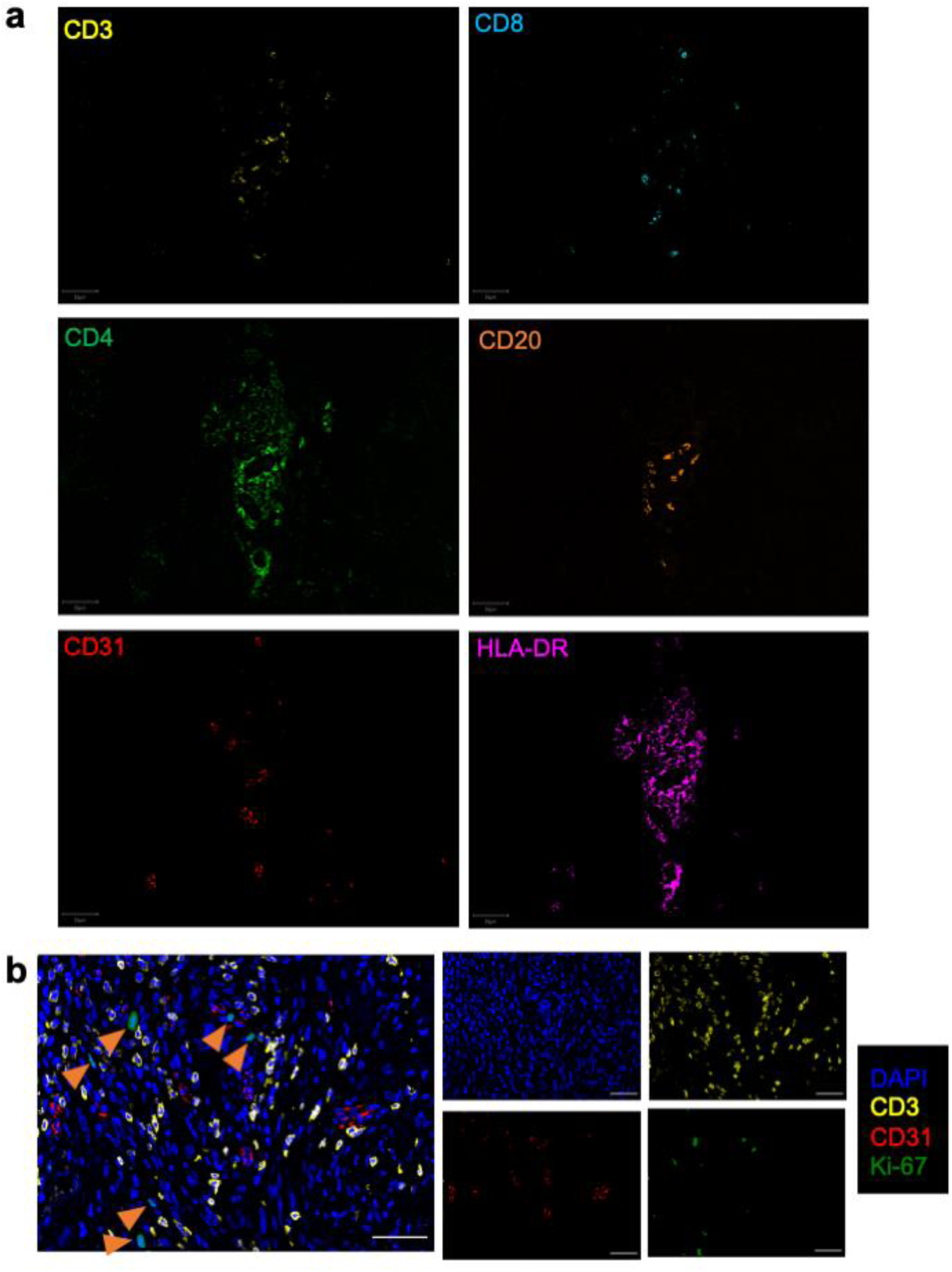
**a**. Single channels of multiplex immunofluorescence images using the PhenoCycler of AAT Material profiling blood vessels (CD31^+^), adaptive immune cells: B cells (CD20^+^), CD3^+^ T Cells (CD4^+^ and CD8^+^), and antigen presenting cells (APCs) (HLA-DR^+^). Scale bar=50 µm **b.** Multiplex immunofluorescence images staining CD3^+^ T cells, CD31^+^ endothelial cells and Ki-67^+^ cell proliferation marker using the PhenoCycler revealed proliferative cells in AAT Material. Scale bar=50 µm

**Supplemental Figure 12.**
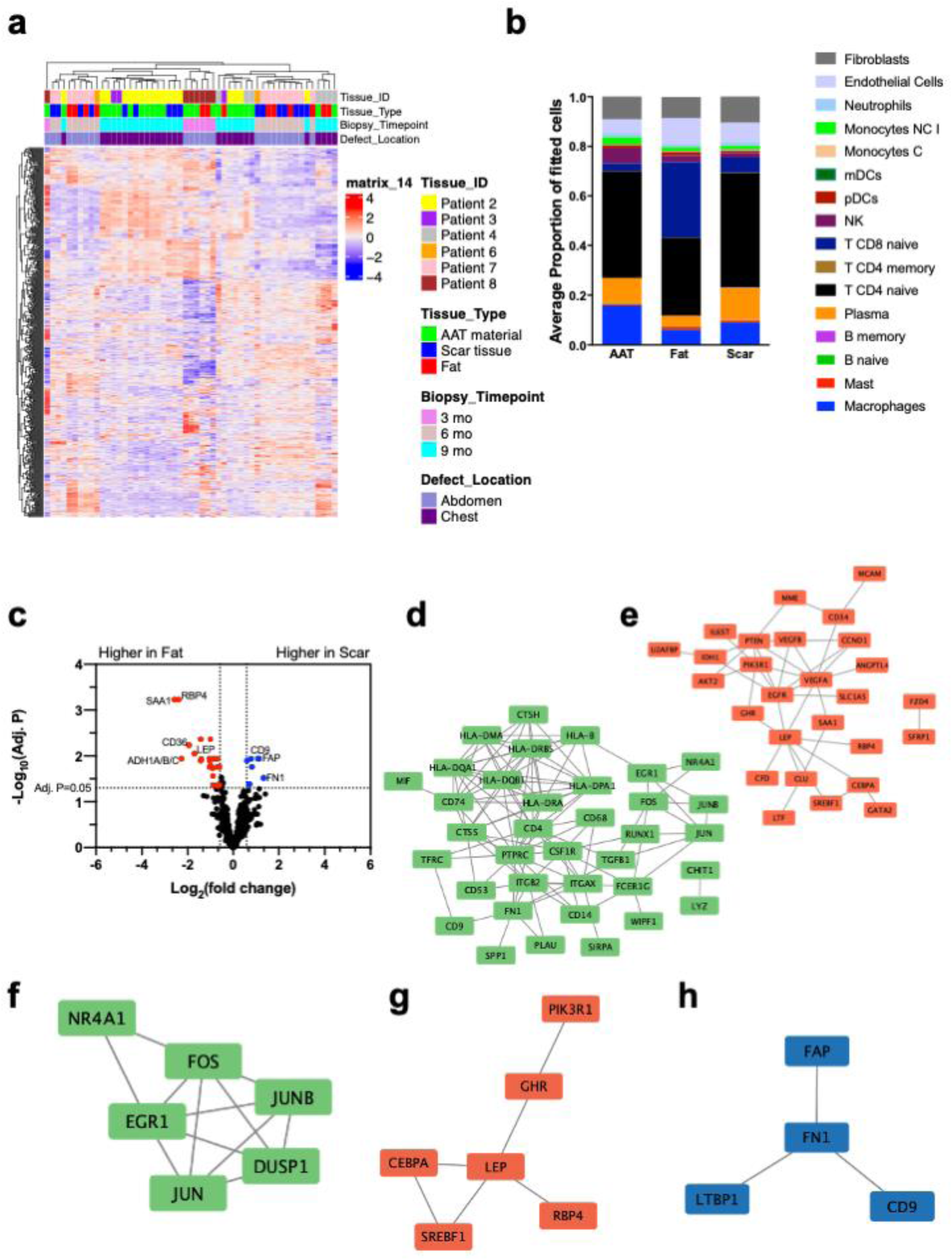
**a**. Hierarchical clustering of all CD31^-^ Nanostring DSP data. Row vst normalized gene counts **b.** Results from cell deconvolution algorithm SpatialDecon applied to CD31^-^Nanostring DSP data represented as average of proportion of fitted cells by tissue type reveal distinct cell composition by tissue type. **c.** Volcano plot of differentially expressed genes comparing fat and scar tissue in core-needle biopsies. Linear mixed model (LMM) test using Limma, design accounts for individual patients, p_adj_ < 0.05, 0.58 ≤ LFC ≤ –0.58 **d.** PPI network of the differentially expressed genes in AAT from AAT and fat comparison, created using the STRING Database; disconnected nodes are not shown for clarity. Edges indicate both known functional and physical protein associations (high confidence-0.7) **e.** PPI network of the differentially expressed genes in fat from AAT and fat comparison, created using the STRING Database; disconnected nodes are not shown for clarity. Edges indicate both known functional and physical protein associations (high confidence-0.7) **f.** PPI network of the differentially expressed genes in AAT from AAT and scar comparison, created using the STRING Database; disconnected nodes are not shown for clarity. Edges indicate both known functional and physical protein associations (high confidence-0.7) **g.** PPI network of the differentially expressed genes in fat from fat and scar comparison, created using the STRING Database; disconnected nodes are not shown for clarity. Edges indicate both known functional and physical protein associations (high confidence-0.7) **h.** PPI network of the differentially expressed genes in scar from fat and scar comparison, created using the STRING Database; disconnected nodes are not shown for clarity. Edges indicate both known functional and physical protein associations (high confidence-0.7)

**Supplemental Figure 13.**
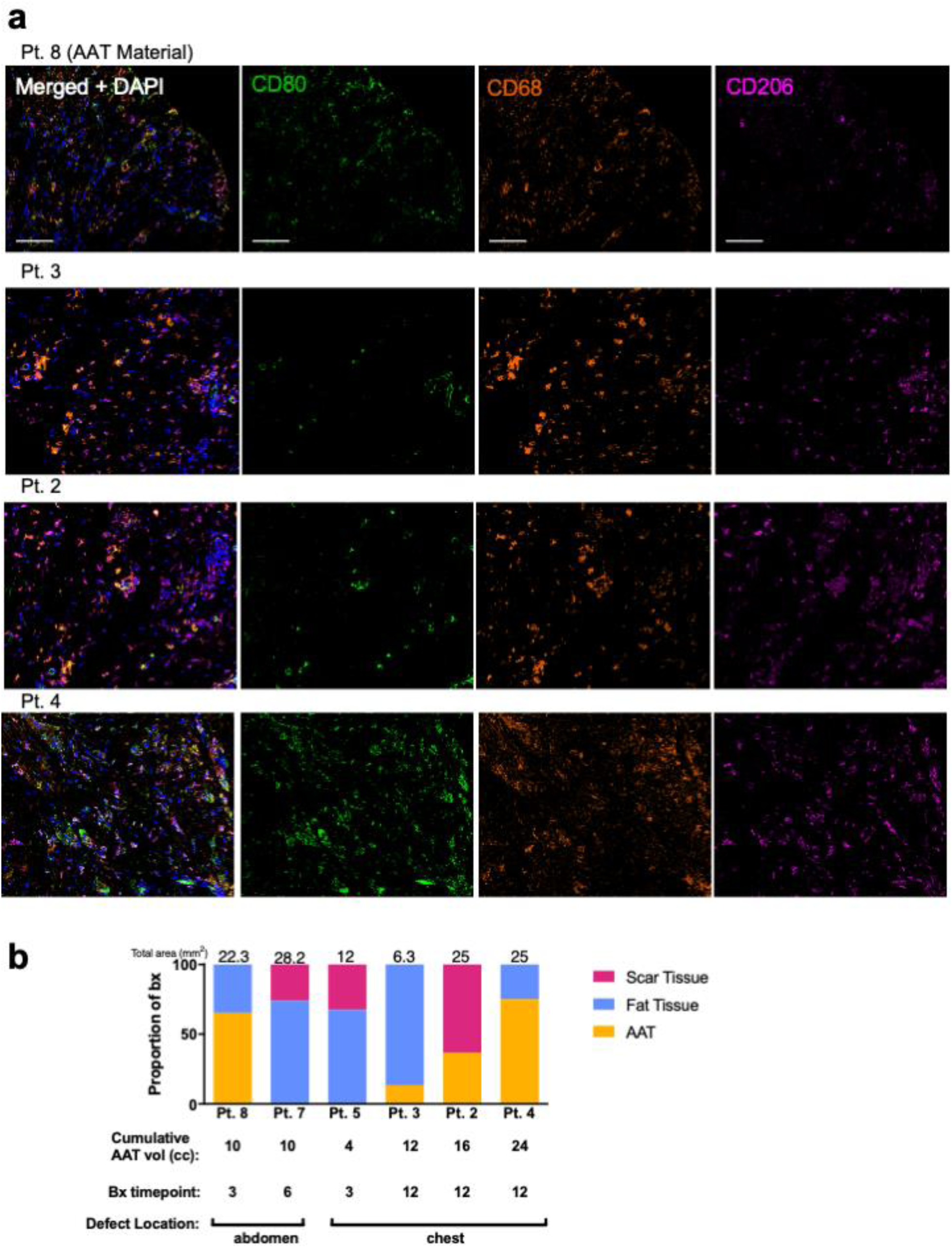
**a**. Single channels for immunofluorescence staining of CD80/CD68/CD206 in AAT material. Scale bar=100 µm **b.** Bar plot of proportion of core-needle biopsy of CD80/CD68/CD206 IF data by tissue type with total area analyzed highlighted

**Supplemental Figure 14.**
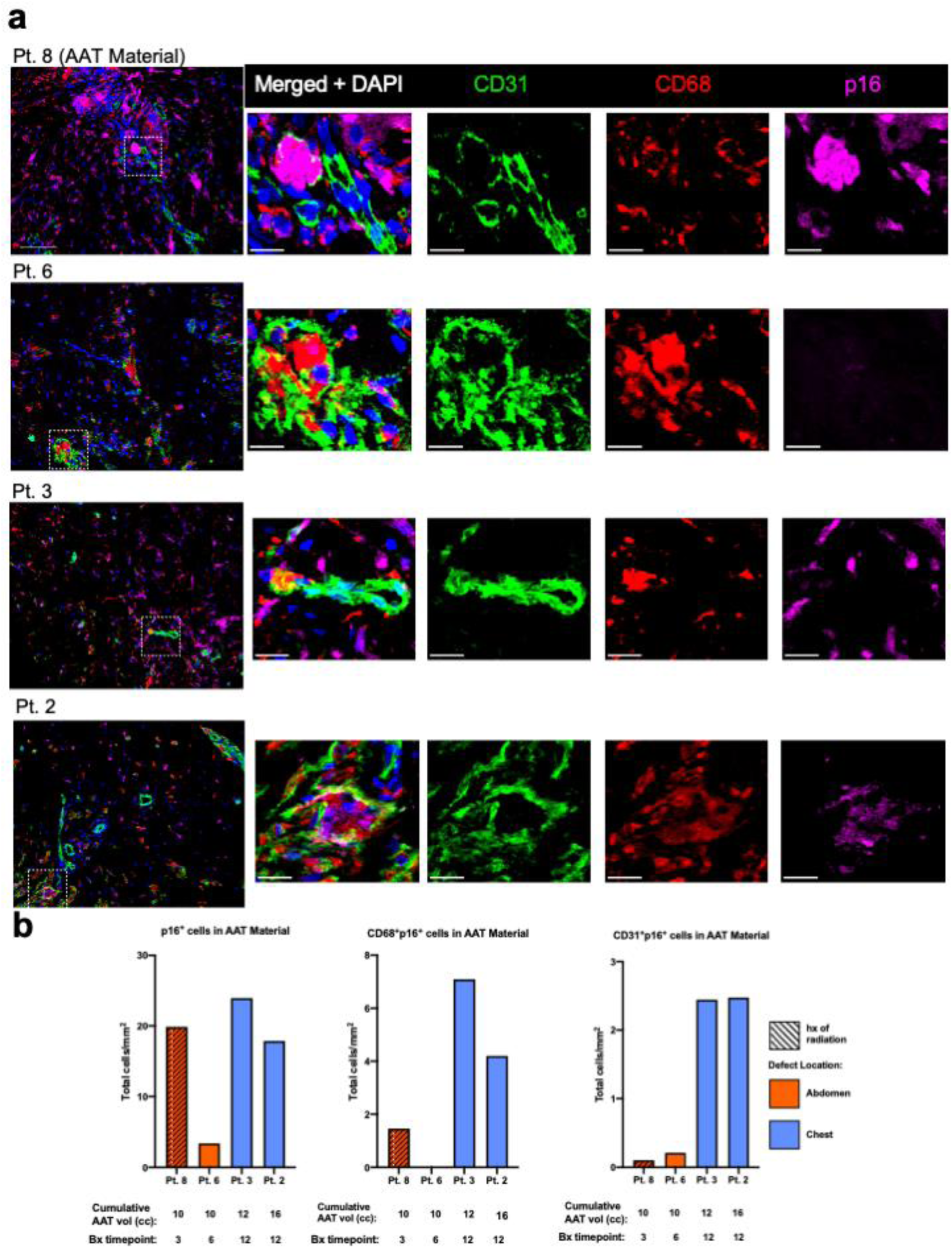
Angiogenesis in AAT Core Needle Biopsies. **a.** Multiplex immunofluorescence staining of endothelial cell (CD31^+^), pan-macrophage (CD68^+^), and senescent marker (p16^+^) reveal macrophages in contact with blood vessels and senescent cell presence in AAT material. Zoomed out image; scale bar=100 µm. Zoomed in image; scale bar=20 µm **b.** p16^+^ cell density, CD68^+^p16^+^ cell density, and CD31^+^p16^+^ cell density in AAT material from IF staining quantified using HALO.

**Supplemental Figure 15.**
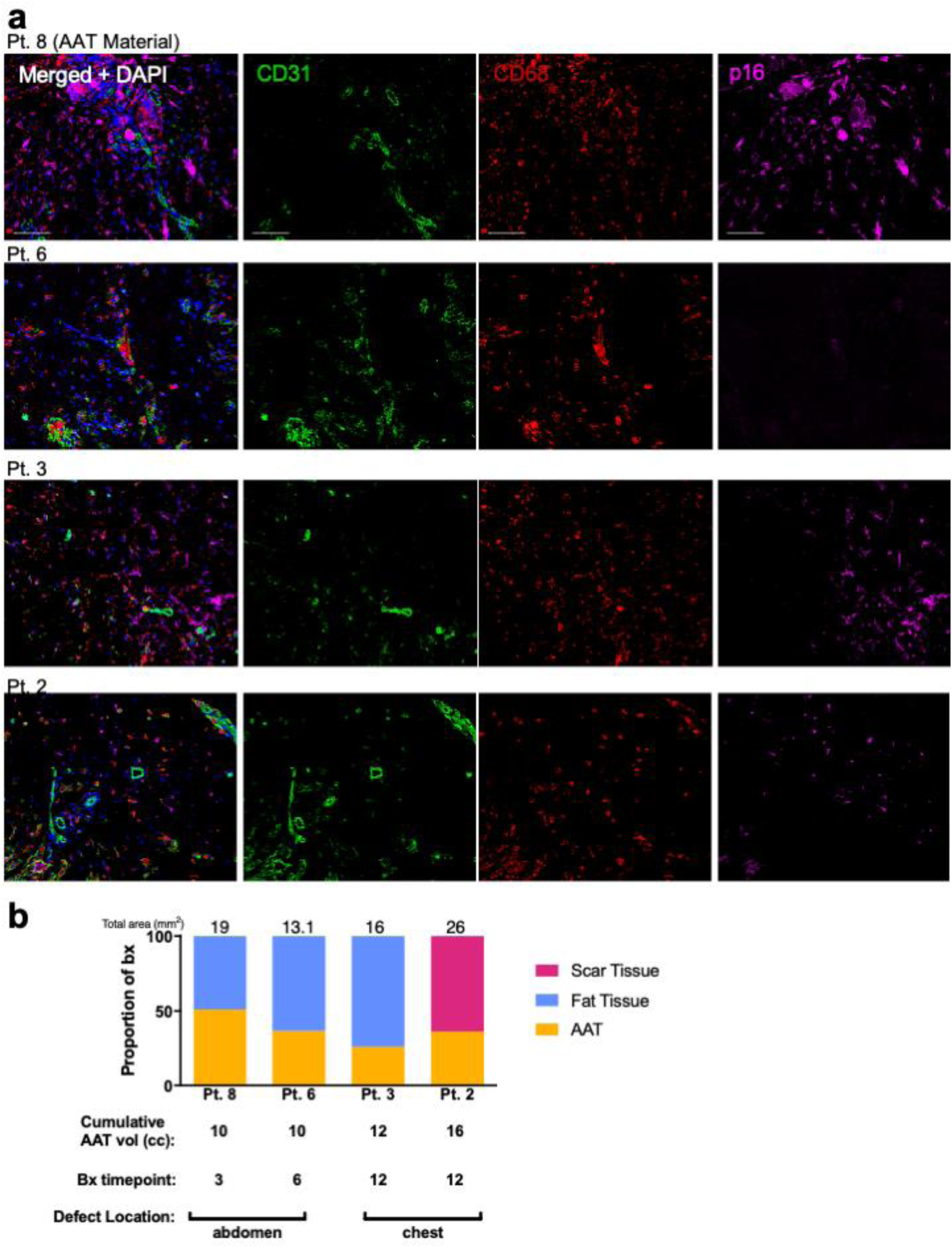
**a**. Single channels for immunofluorescence staining of CD31/CD68/p16 in AAT material. Scale bar 100 µm **b.** Bar plot of proportion of core-needle biopsy of CD31/CD68/p16 IF data by tissue type with total area analyzed highlighted

**Supplemental Figure 16.**
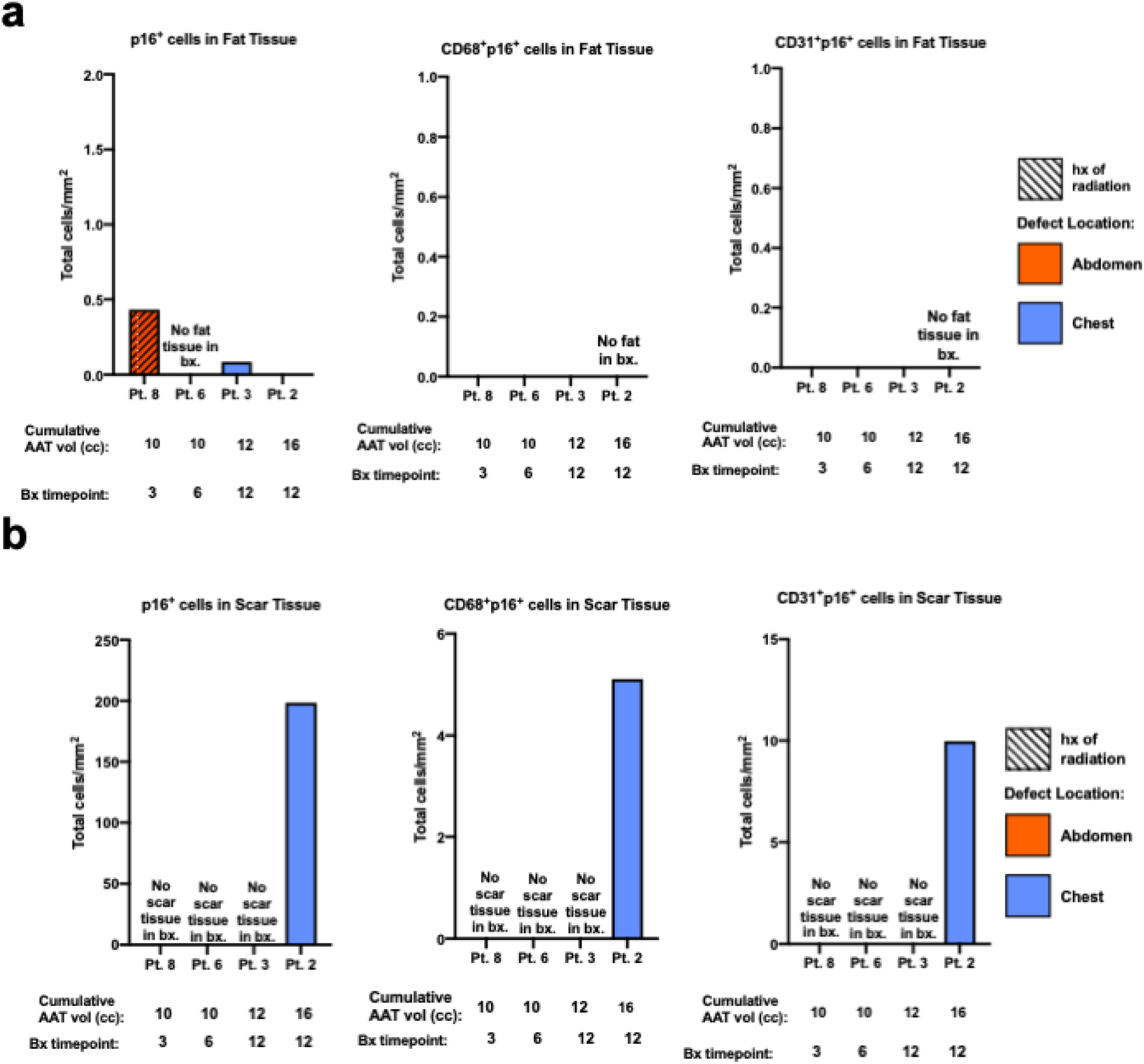
**a**. p16^+^ cell density, CD68^+^p16^+^ cell density, and CD31^+^p16^+^ cell density in fat tissue from IF staining quantified using HALO. **b.** p16^+^ cell density, CD68^+^p16^+^ cell density, and CD31^+^p16^+^ cell density in scar tissue l from IF staining quantified using HALO.

**Supplemental Figure 17.**
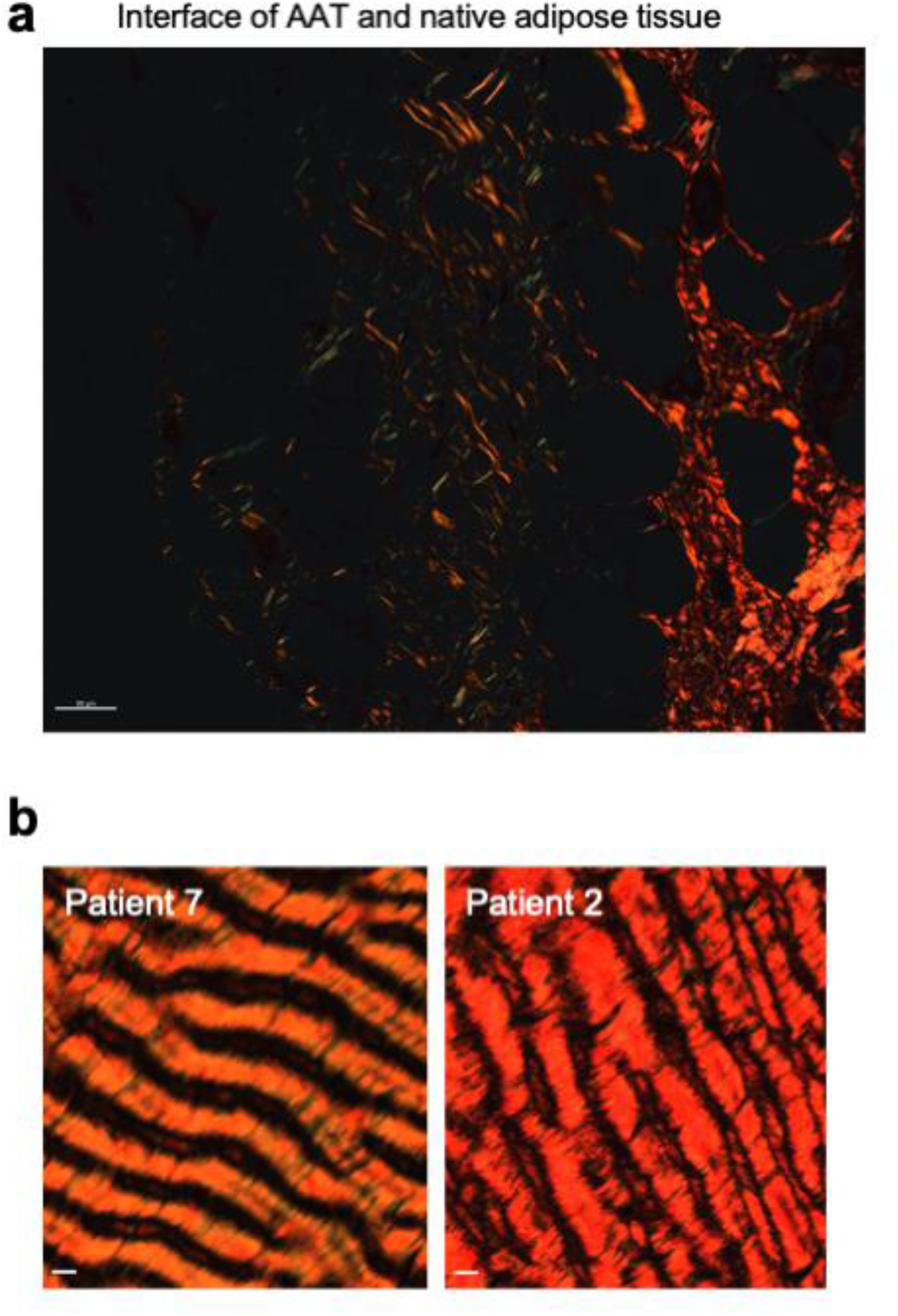
Additional PSR images of core-needle biopsies. **a.** Polarized light images of PSR staining at AAT native adipose tissue interface. Scale bar=50 µm **b.** Polarized light images of PSR staining of scar tissue to investigate collagen network remodeling. Scale bar=10 µm

**Supplemental Figure 18.**
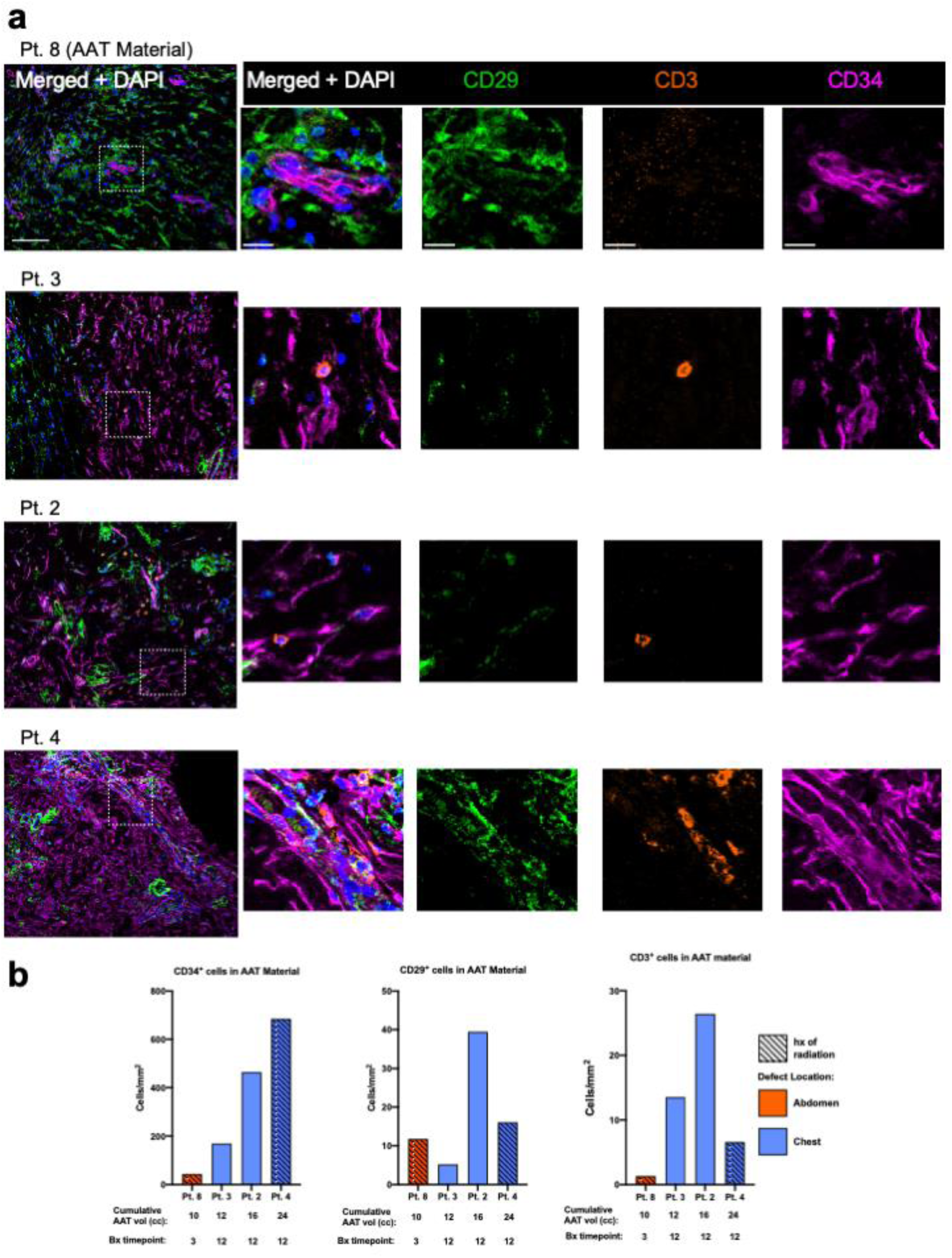
Stromal, progenitor, and T cell profiling of AAT core-needle biopsies. **a.** Multiplex immunofluorescence staining of stromal (CD29^+^), T cell (CD3^+^), and progenitor (CD34^+^) marker revealed presence of these cell types and cell-cell interactions. Zoomed out image; scale bar=100 µm. Zoomed in image; scale bar=20 µm **b.** CD34^+^ cell density, CD29^+^ cell density, and CD3^+^ cell density in AAT material quantified by IF staining using HALO

**Supplemental Figure 19.**
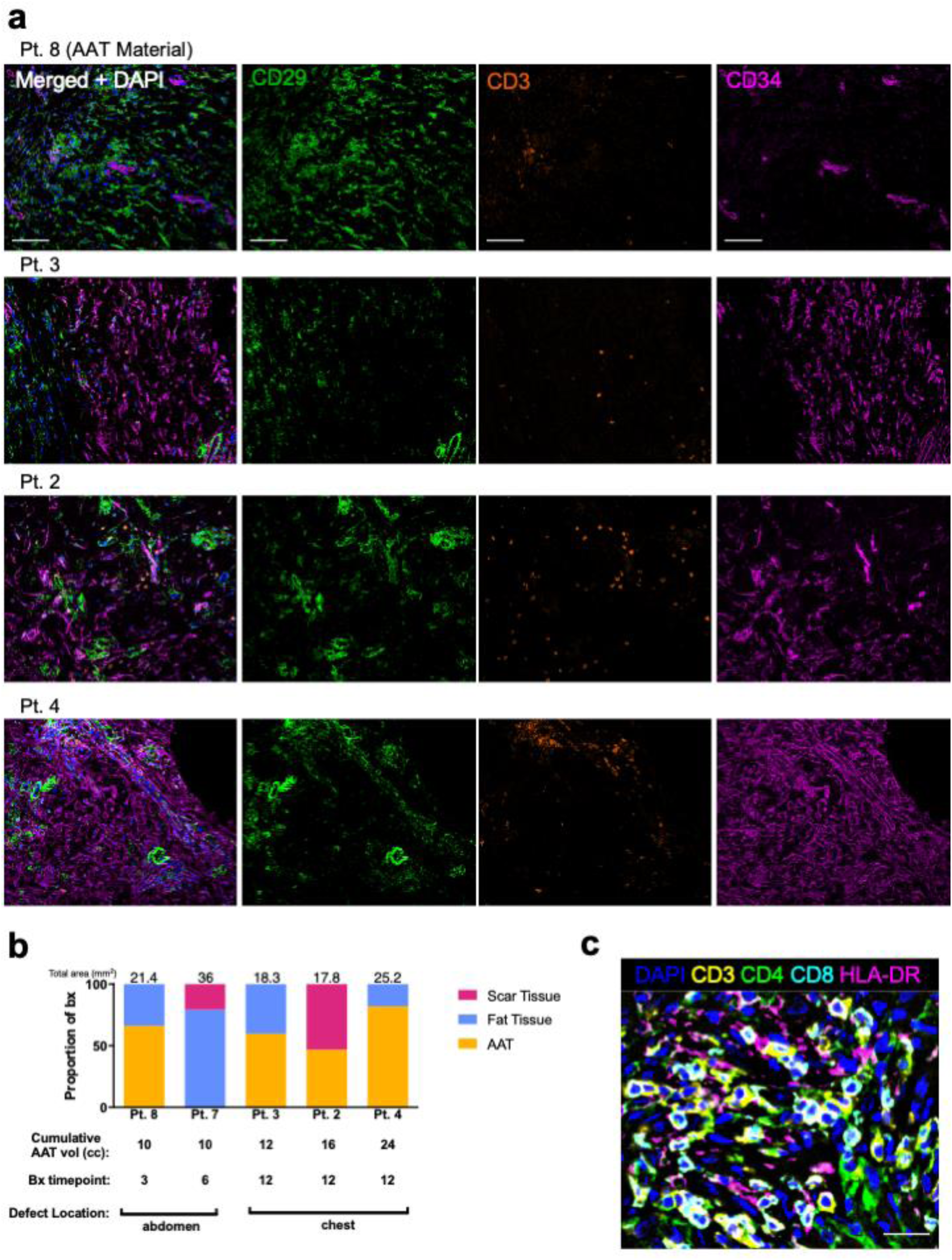
**a**. Single channels for immunofluorescence staining of CD29/CD3/CD34 in AAT material. Scale bar=100 µm **b.** Bar plot of proportion of core-needle biopsy of CD29/CD3/CD34 IF data by tissue type with total area analyzed highlighted **c.** Multiplex immunofluorescence images staining CD3^+^CD4^+^ T cells, CD3^+^CD8^+^ T cells in contact with HLA-DR^+^ antigen presenting cells using the PhenoCycler in AAT Material. Scale bar= 20 µm

## Notes

### Clinical Trial

NCT03544632

### Author Declarations

IRB– and OHRO– (formerly known as the HRPO)-approved Phase II clinical trial at Johns Hopkins Medicine (Baltimore, MD) IRB (Study ID# IRB00155003) and the OHRO Log No. E01422.1a.

