## Supplemental Tables for "Acellular Adipose Tissue promotes anti-fibrotic remodeling in Phase II Study"

Supplemental Table 1 Patient information

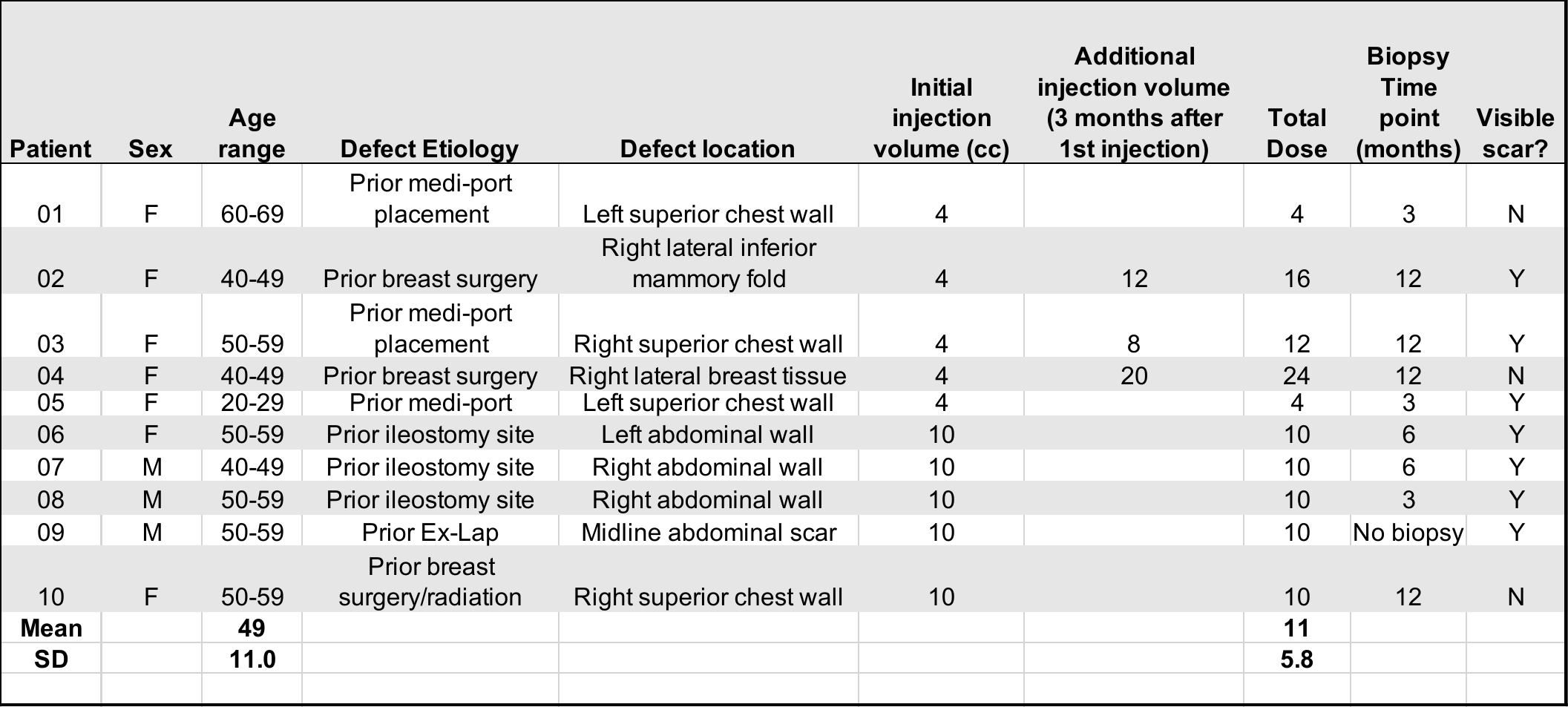

Supplemental Table 2 Patient gender and race overview

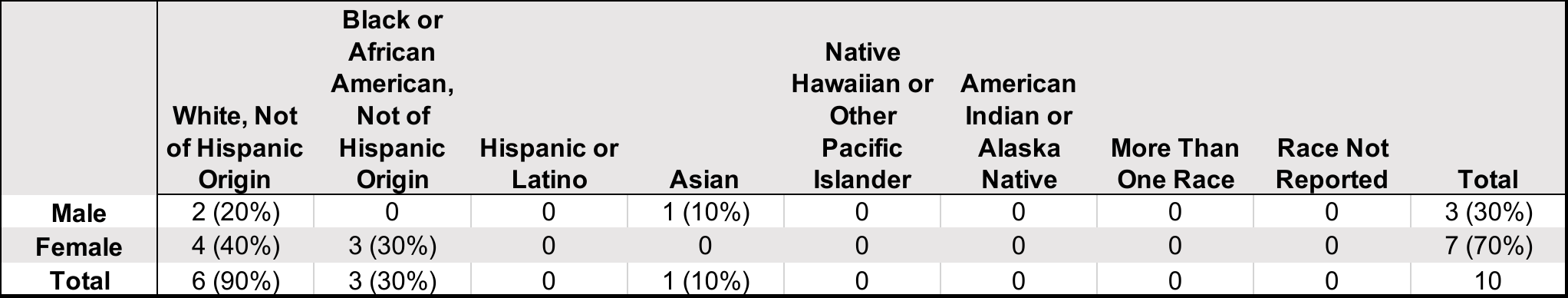

Supplemental Table 3 Patient soft tissue defect summary

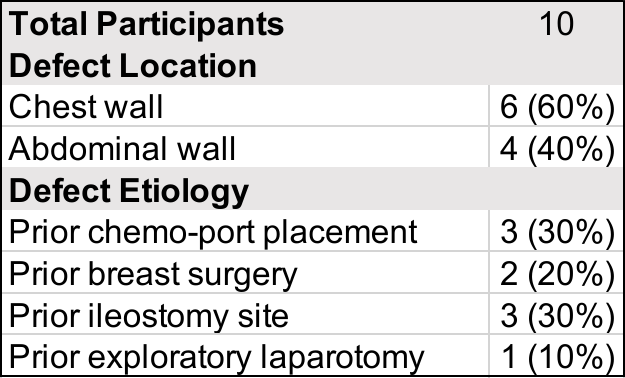

Supplemental Table 4 Overview of physical exam findings

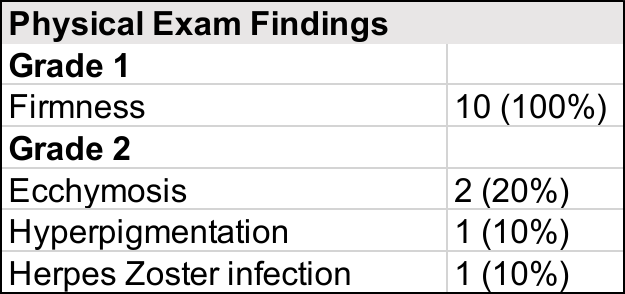

Supplemental Table 5 Anticipated Adverse Events Related to Injection Sites

| **Anticipated AEs** | |
| --- | --- |
| Pain/Tenderness | 1 (10%) |
| Erythema | 1 (10%) |
| Bruising | 1 (10%) |
| Hyperpigmentation | 1 (10%) |
| Textural change | 0 |

Supplemental Table 6 Summary of Adverse Events by System Organ Class (n=10)

| **ASAE Body System** | **Adverse Event** | **# of Occurrences** | **# of Subjects** | **Grade** | **Relatedness to AAT** | **Outcome** |
| --- | --- | --- | --- | --- | --- | --- |
| Skin and subcutaneous tissue disorders | Skin hyperpigmentation | 1 | 1 | 1 | Probably Related | Resolved |
| Skin and subcutaneous tissue disorders | Ecchymosis or Bruising | 2 | 2 | 1 | Probably Related | Resolved |
| Infections and infestations | Shingles | 1 | 1 | 2 | Probably Not Related | Resolving |
| Gastrointestinal disorders | Dry mouth | 2 | 2 | 1 | Not Related | Resolved |
| Gastrointestinal disorders | Recurrence of prior abdominal wall hernia | 1 | 2 | 2 | Not Related | Ongoing |
| General disorders and administration site conditions | Fatigue | 1 | 1 | 1 | Probably Not Related | Resolved |
| Investigations | Elevated Alanine Aminotransferase | 1 | 1 | 1 | Probably Not Related | Resolved |
| Investigations | Elevated Aspartate Aminotransferase | 1 | 1 | 1 | Probably Not Related | Resolved |
| Surgical and medical procedures, Other, specify | Recurrence of prior abdominal wall hernia | 1 | 1 | 1 | Probably Not Related | Ongoing |
| Infections and Infestations | Urinalysis concerning for urinary tract infection | 1 | 1 | 2 | Not Related | Resolved |
| Investigations | Elevated C-reactive protein | 4 | 3 | 1 | Possibly Related | Resolved |

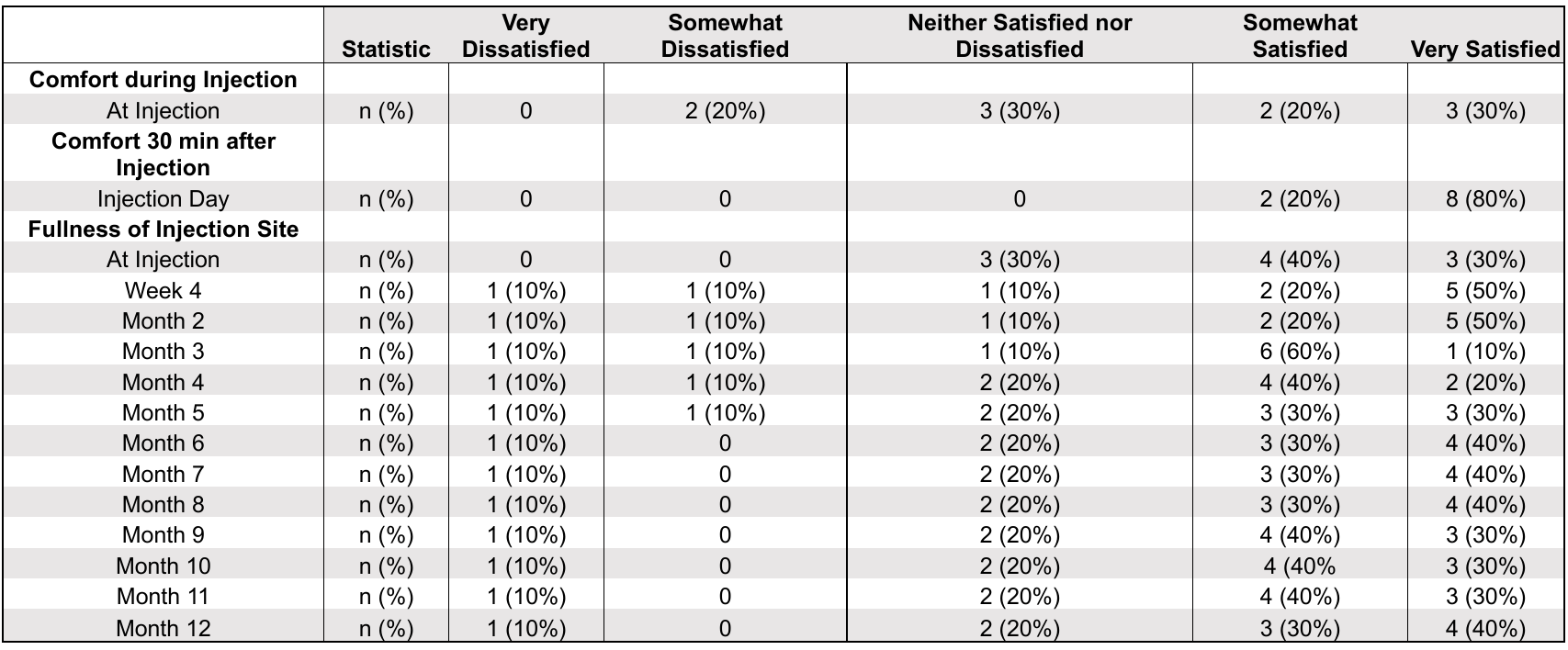

Supplemental Table 7 Summary of Participant Survey results for comfortability and fullness

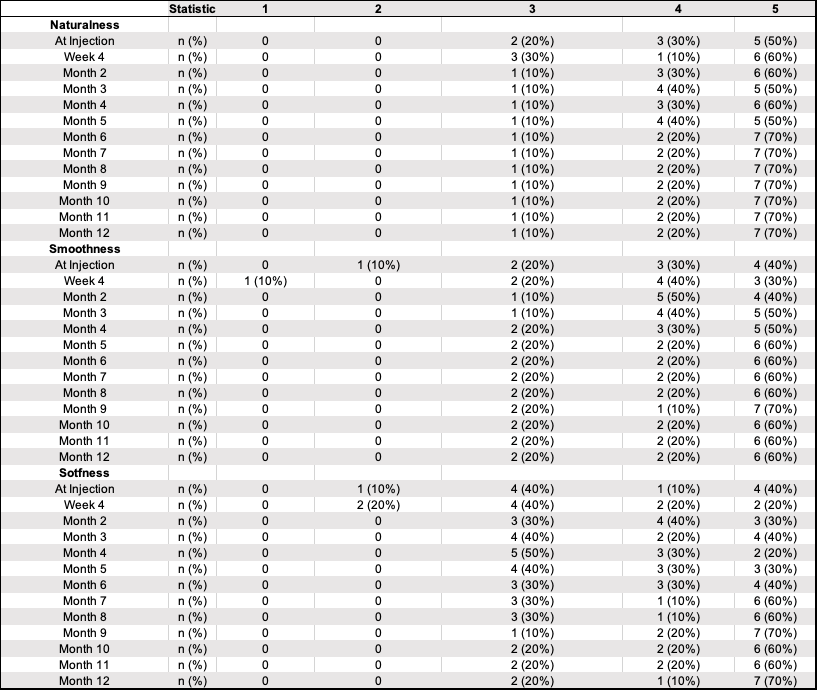

Supplemental Table 8 . Summary of Participant Survey results for naturalness, smoothness, and softness

Supplemental Table 9 Summary of Investigator Ease of Use Surveys for each subject (n=10)

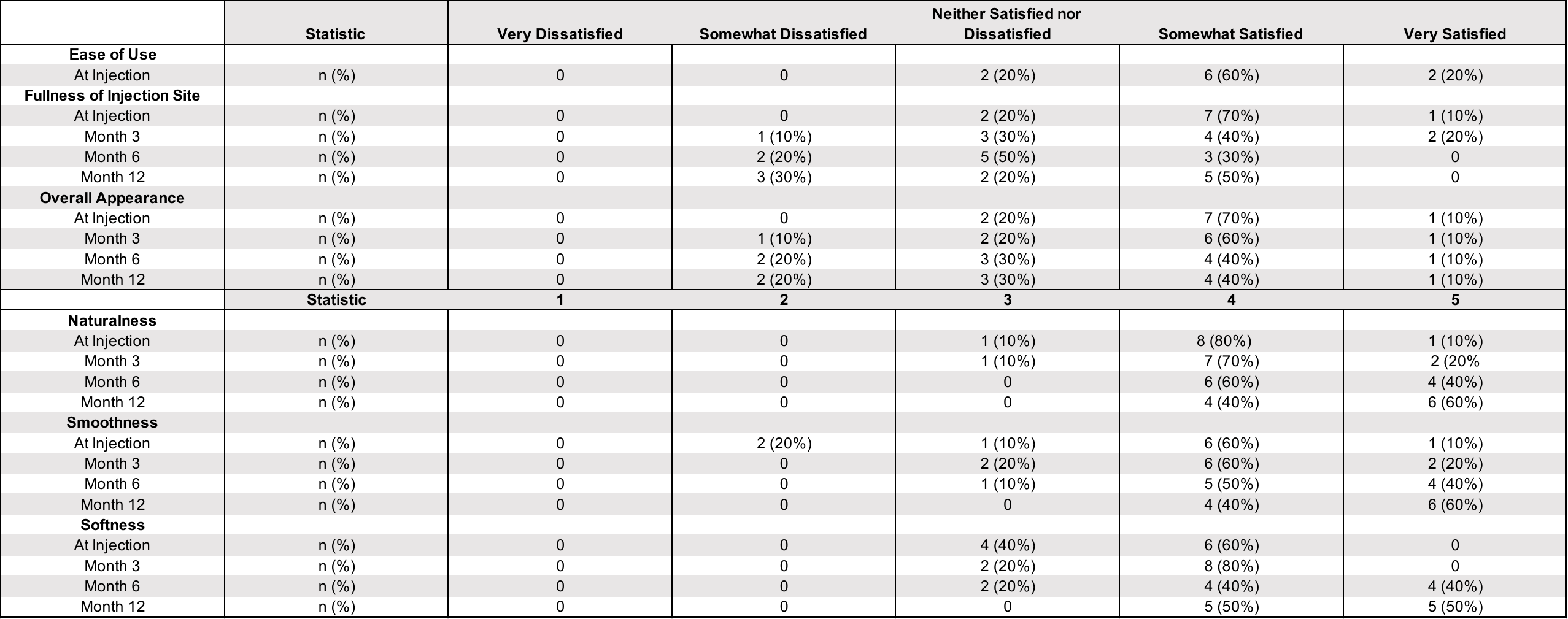

Supplemental Table 10 Panel Reactive Testing Results

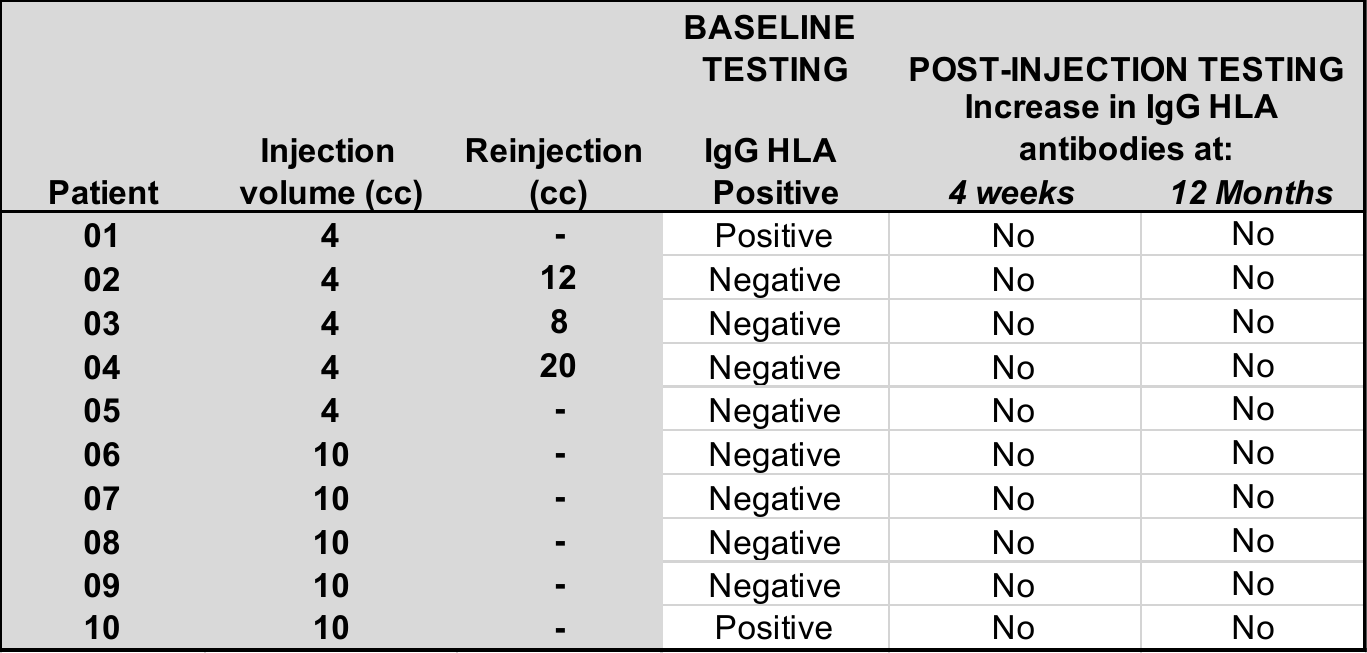

Supplemental Table 11 Available interval CT scan volume measurements

| **Patient** | **Injection** | **Timepoint** | **CT Scan**  **Volume** | **CT % Retained** |
| --- | --- | --- | --- | --- |
| **06** | 10cc | 3 mo | 5.267cc | 52.7% |
| **08** | 10cc | 2 mo | 5.345cc | 53.5% |

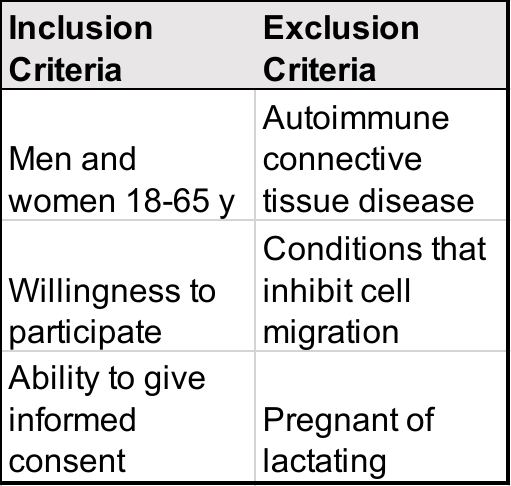

Supplemental Table 12 Inclusion and exclusion criteria

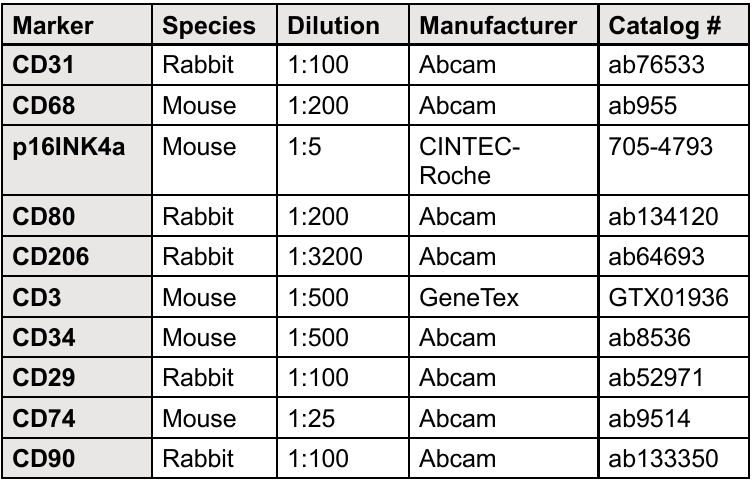

Supplemental Table 13 Antibodies used in Opal/TSA immunofluorescence staining

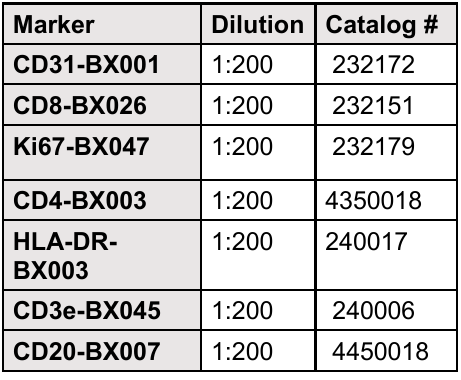

Supplemental Table 14 Antibodies used in CODEX staining panel
